# Aging-related cell type-specific pathophysiologic immune responses that exacerbate disease severity in aged COVID-19 patients

**DOI:** 10.1101/2021.09.13.21263504

**Authors:** Yuan Hou, Yadi Zhou, Michaela U. Gack, Yuan Luo, Lara Jehi, Timothy Chan, Haiyuan Yu, Charis Eng, Andrew A. Pieper, Feixiong Cheng

**Author notes:** Correspondence to: Feixiong Cheng, PhD, Lerner Research Institute, Cleveland Clinic.

## Abstract

Coronavirus Disease 2019 (COVID-19) is especially severe in aged patients, defined as 65 years or older, for reasons that are currently unknown. To investigate the underlying basis for this vulnerability, we performed multimodal data analyses on immunity, inflammation, and COVID-19 incidence and severity as a function of age. Our analysis leveraged age-specific COVID-19 mortality and laboratory testing from a large COVID-19 registry, along with epidemiological data of ∼3.4 million individuals, large-scale deep immune cell profiling data, and single-cell RNA-sequencing data from aged COVID-19 patients across diverse populations. To begin, we confirmed a significantly increased rate of severe outcomes in aged COVID-19 patients. Furthermore, we identified increased inflammatory markers (C-reactive protein, D-dimer, and neutrophil-lymphocyte ratio), viral entry factors in secretory cells, and TGFβ-mediated immune-epithelial cell interactions, as well as reduction in both naïve CD8 T cells and expression of interferon antiviral defense genes (*i*.*e*., *IFITM3* and *TRIM22*), along with strong TGF-beta mediated immune-epithelial cell interactions (i.e., secretory - T regulatory cells), in aged severe COVID-19 patients. Taken together, our findings point to immuno-inflammatory factors that could be targeted therapeutically to reduce morbidity and mortality in aged COVID-19 patients.

## Introduction

Coronavirus Disease 2019 (COVID-19), a global pandemic caused by severe acute respiratory syndrome coronavirus 2 (SARS-COV-2), has been diagnosed in more than 170 million people globally, with 3.5 million deaths since December 2019 (data on May 31, 2021). Although a serious risk at any age, SARS-CoV-2 infection is particularly debilitating and deadly for aged patients, defined in this study as 65 years and older^1,2,3,4,5^. The molecular basis of this aging-related vulnerability is an important area of investigation as it is currently poorly understood.

Impaired and dysregulated host immunity, including both innate and adaptive immunity, have been hypothesized as age-based factors in COVID-19 disease severity^4,6^. Compared to younger individuals with COVID-19, aged individuals show disrupted antigen-specific adaptive immunity to SARS-CoV-2, such as reduced coordination of CD4-CD8 T cell responses^7^. In addition, aged individuals typically produce a less robust type I interferon (IFN) response to flu virus infections^8^, indicating compromised cellular antiviral defense in innate immunity. Indeed, 13% of aged patients with life-threatening COVID-19 display inborn errors in autoantibodies against type I IFN immunity^9^. In addition, aberrant immunosenescence and inflammation also play crucial roles in age-medicated COVID-19 morbidity and mortality^10^. For example, senescent cells become hyper-inflammatory in response to pathogen-associated molecular patterns, and senolytics reduce COVID-19-mortality in aged mice^11^. Based on these findings, we sought to systematically identify whether there are specific immuno-inflammatory determinants that promote age-associated COVID-19 severity.

## Results

### Severe outcomes in aged COVID-19 patients

To begin, we investigated the prevalence of COVID-19 disease among different age groups with nine months data collection. Analysis of U.S. Centers for Disease Control (CDC) epidemiological data from March to December, 2020 (**Supplementary Tables 1-3**) revealed that 80.5% of fatal-cases occurred in aged patients. Strikingly, this rate was 4.1 times higher than in 18-64 years old (19.5%), and 1,653 times higher than in 0-17 years old (0.05%, **Fig. 1a**). Fatality prevalence was influenced by sex in both older and younger groups (**Fig. 1b**). Notably, the relative fatality prevalence trend across these same three age groups from 2010-2020 with influenza virus was similar, (**Fig. 1a**), indicating that age is a universal risk factor for viral infections, especially for older males.

**Figure 1.**
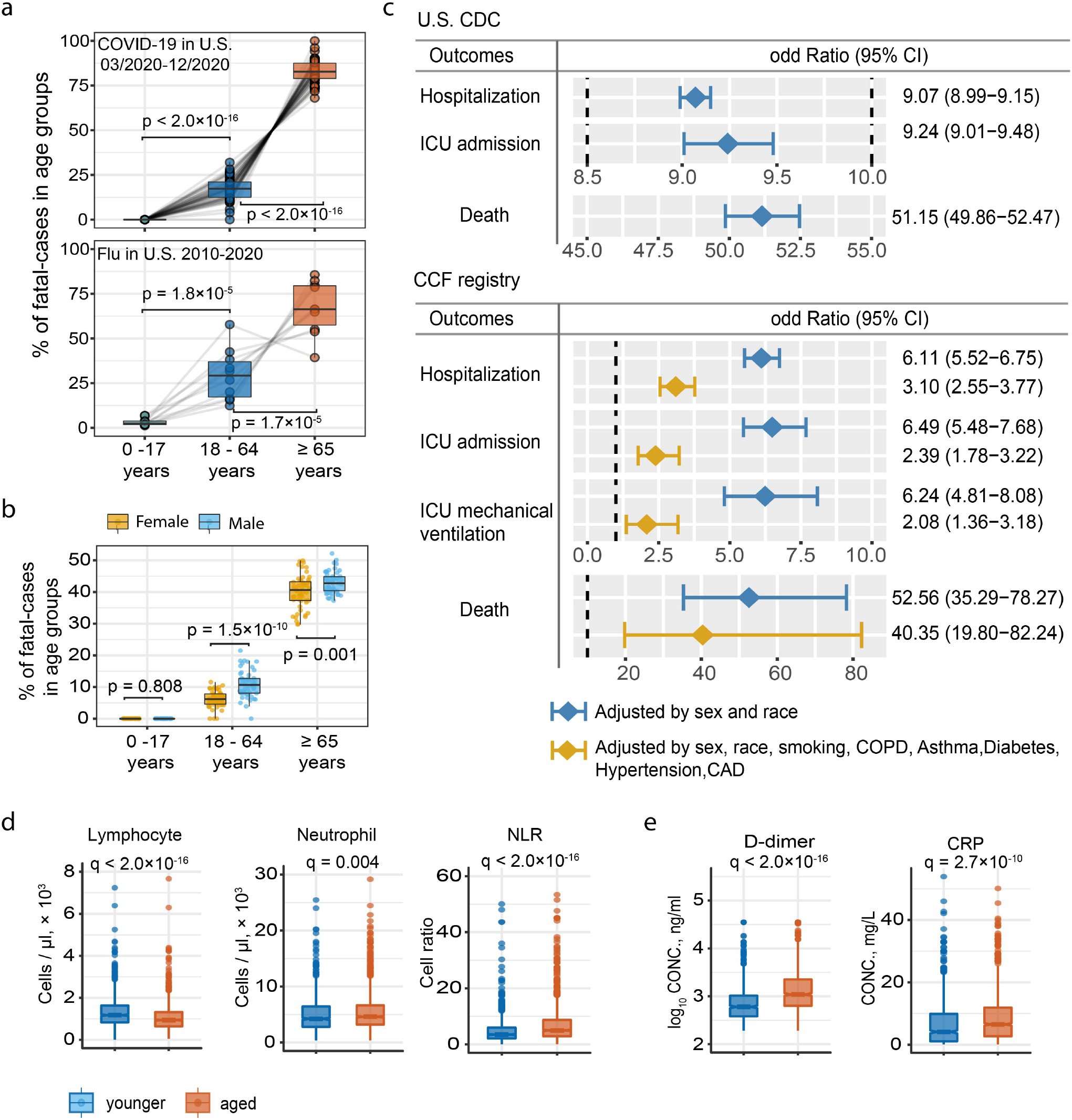
Epidemiological data analysis between aged and younger COVID-19 patients. **a**, The percentage of fatal-cases of COVID-19 and flu across three age groups. Data source from United States (U.S.) CDC. The upper panel shows the percentage of fatal-cases of COVID-19 in U.S. Each dot in the boxplot represents one state. The lower panel shows the percentage of fatal-cases of flu from 2010-2020. Each dot in the boxplot represent one flu season. Statistical p-value was computed by two tailed paired t-test. **b**, Sex differences in the percentage of fatal-cases of COVID-19 across three age groups. **c**, Odds ratio (OR) analysis of U.S. CDC and COVID-19 registry datasets. U.S. CDC dataset. **d** and **e** Boxplot show the lab testing values of four inflammatory markers between aged and younger individuals. Adjusted p-value [q] was computed by Mann–Whitney *U* test with Benjamini–Hochberg (BH) multiple testing correction.

Next, we used odds ratio (OR) adjusted for confounding factors to further evaluate the association between aging and four types of COVID-19 outcomes: hospitalization, intensive care unit (ICU) admission, and death. Specifically, we analyzed sex- and race-adjusted OR values in 3,417,930 COVID-19 positive cases (n = 2,369,919 in young individuals, 20-49 years old) and n = 1,048,011 in aged individuals (> 60 years old); see Method, **Supplementary Table 3**) from the U.S. CDC database. Here, aged individuals showed significantly increased likelihood of COVID-19-related hospitalization (OR = 9.07, 95% confidence interval [CI] 9.99 – 9.15, **Fig. 1c**), ICU admission (OR = 9.24, 95% CI 9.01 – 9.48), and death (51.15, 95% CI 49.86 – 52.47, **Fig. 1c**).

To further account for disease comorbidities, we next computed OR across different age groups using a large COVID-19 registry database with 12,651 aged (≥ 65 years) and 32,426 younger individuals (20 - 55 years old) (**Fig. 1c, Supplementary Table 4**, see Methods). Specifically, we tested the OR Model-2, which is adjusted for sex, race, smoking, and five common disease comorbidities^12,13^ (hypertension, diabetes, coronary artery disease, asthma, chronic obstructive pulmonary disease [COPD] & emphysema). Here, we again found that aged individuals had significantly greater likelihood of COVID-19-related hospitalization (OR = 3.10, 95% CI 2.55 – 3.77), ICU admission (OR = 2.39, 95% CI 1.78 – 3.22) (**Fig. 1c**), and death (OR = 40.35, 95% CI 19.80 – 82.24). Subsequent Kaplan-Meier analysis further revealed an elevated cumulative hazard for hospitalization (p < 0.0001, Log-rank test, **Supplementary Fig 1a**), including longer duration of hospitalization (average duration = 8.9 days; p = 1.4 ×10^−15^, Mann–Whitney *U* test, **Supplementary Fig 1b**), in COVID-19 patients. Taken together, our findings confirm an elevated likelihood of severe outcomes in aged COVID-19 patients have compared to younger patients, even when adjusted for all possible confounding factors.

### Elevated inflammatory responses in aged COVID-19 patients

As severe COVID-19 patients have been reported to have lower lymphocyte count^14^ and higher C-reactive protein (CRP)^15^, we examined the Cleveland Clinic COVID-19 registry for differences in inflammatory biomarkers as a function of aging. Here, we found lower peripheral lymphocytes (adjusted p value [q] < 2.0 × 10^−16^, Mann–Whitney *U* test with Benjamini-Hochberg multiple test correction, **Fig. 1d**) and higher circulating neutrophils in hospitalized aged COVID-19 patients (q =0.004, **Fig. 1d**), compared to younger patients. We also found that the neutrophil-lymphocyte ratio (NLR), a marker of systemic inflammation^16^, was elevated in aged COVID-19 patients (q < 2.0 × 10^−16^, **Fig. 1d**). In addition, the inflammatory markers D-dimer (q < 2.0 × 10^−16^, **Fig 1e**) and C-reactive peptide (CRP) (q = 2.7 × 10^−10^, **Fig 1e**) were also significantly increased in hospitalized aged compared to hospitalized young COVID-19 patients. Taken together, our findings identify elevated inflammatory response and decreased lymphocyte count as potential mediators of severe COVID-19 outcomes in aged patients.

### Elevated pro-inflammatory cytokine expression in aged COVID-19 patients

We next examined peripheral immune profiles^17^ of hospitalized aged and younger COVID-19 patients by querying a dataset of 12 major immune cell types (% peripheral blood mononuclear cells [PBMCs]) and 32 T cell subtypes (% CD3, **Supplementary Table 5**, see Methods). There was no difference in abundance of the major immune cell types (*e*.*g*. T cells, B cells, natural killer cells, and plasmacytoid dendritic cells [pDC]) between aged and young hospitalized COVID-19 patients, including those in the ICU (**Fig. 2a, c** and **Supplementary Fig. 2a**). However, both young and aged COVID-19 patients with ICU admission had a lower proportion of T cells (younger, q = 0.001; older, q=0.003) and pDC (younger, q = 0.009; older, q=0.004) (**Fig. 2a, c**), as well as an elevated proportion of non-classic Monocytes (ncMono) (younger, q = 0.003; older, q=0.014, **Fig. 2c**), compared to non-ICU patients. Further analysis of deep phenotyping T cell data revealed significantly fewer naïve CD8 T cells in hospitalized aged COVID-19 patients (q = 1.7 × 10^−11^, **Fig. 2b, d**). Naïve CD8 T cell-mediated homeostasis is an important component of antiviral defense^18^, and the naïve CD8 T cell receptor repertoire negatively correlated with age in COVID-19 patients^19^. Reduced abundance of naïve CD8 T cells could dysregulate CD8 T cell pool homeostasis and perturb immunity.

**Figure 2.**
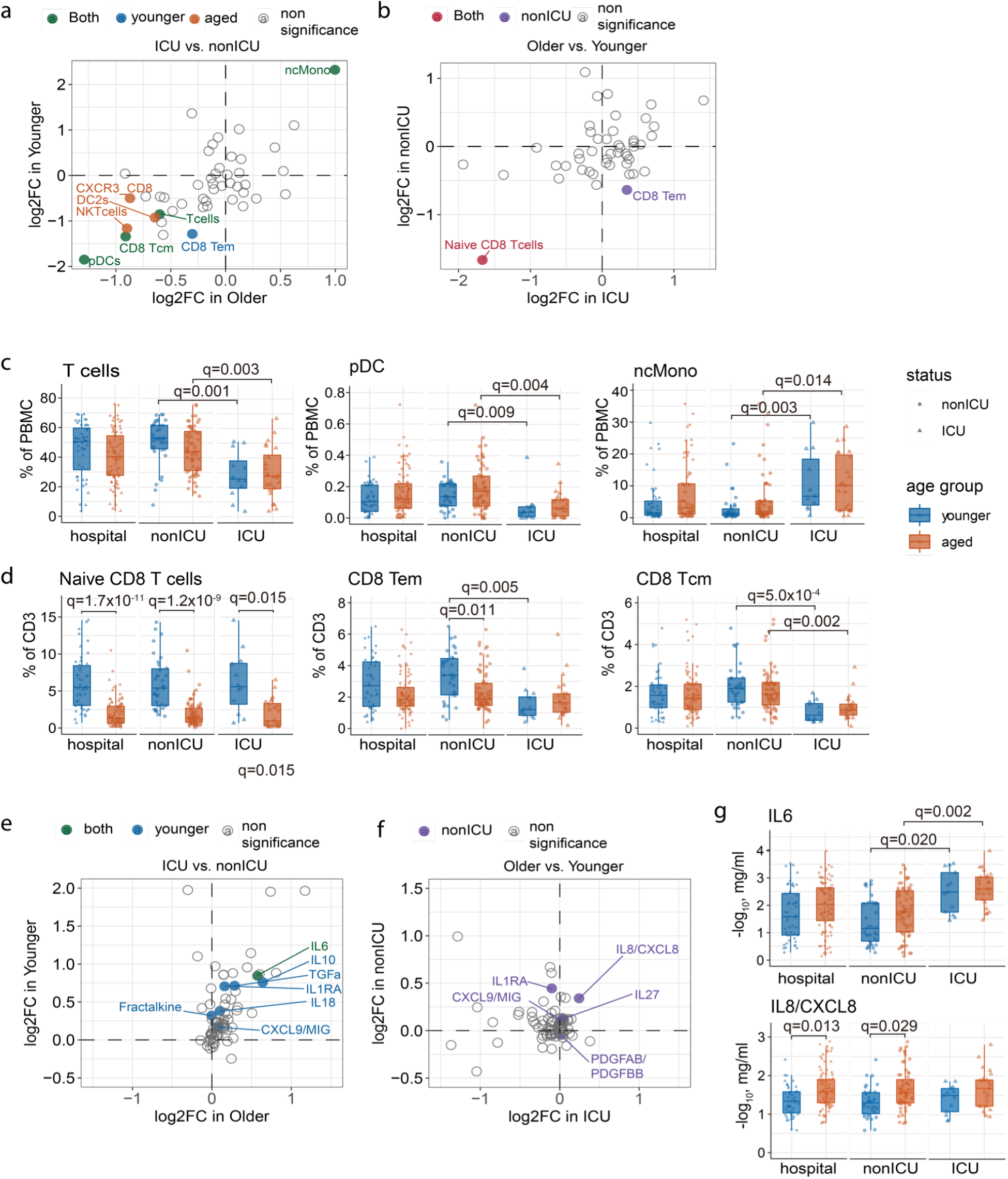
Deep immune-profiling of aged and younger patients with COVID-19. **a** and **e** Scatterplot shows the differential immune cell type (**a**) and cytokines (**e**) between ICU (n = 39 samples) versus non-ICU (105 samples) COVID-19 patients. **b and f**, Scatterplot shows the differential immune cell type (d) and cytokines (h) in aged (n = 94 samples) versus younger (50 samples) COVID-19 patients. **c**, The abundance of major immune cell types in PBMC and (**d**) subtypes of CD8+ T cells in all CD3 positive cells. **g**, The abundance of IL6 and IL8 change between younger and aged COVID-19 patients in hospital, ICU and non-ICU groups.

Next, we compared the plasma profile of 71 cytokines and chemokines^17^ between hospitalized aged and younger COVID-19 patients (**Supplementary Table 5**). Historically, increased IL-6, IL-8, IL-10 and IL-27 levels have been associated with severe COVID-19^20,21^. Here, we found elevated IL-8 (also named CXCL8) and IL-27 expression in aged COVID-19 patients (q = 0.013, **Fig. 2g** and **Supplementary Fig. 2b**). As IL-8 is a pro-inflammatory cytokine that recruits and activates neutrophils^22^, its elevation is consistent with our previously noted elevated neutrophil count and NLR in hospitalized aged COVID-19 patients (**Fig. 1d**). Furthermore, younger, but not aged, COVID-19 ICU patients also showed elevated IL-10 **(Supplementary Fig. 2**), a key feature of cytokine storm^23,24^. In addition, elevated IL-6 was observed in both younger (q = 0.020) and aged ICU patients, (q=0.002), compared to non-ICU patients (**Fig. 2g**). Thus, severe COVID-19 patients show distinct age-related cytokine profiles, with hospitalized aged patients showing elevated IL-8 and IL-27, and hospitalized younger patients showing elevated IL-6 and IL-10. These results indicate that heterogeneous inflammatory cytokine expression between aged and younger COVID-19 patients may mediate age-related hospitalization and ICU admission.

### Reduced naïve CD8 T cells in aged severe COVID-19 patients

Because we observed loss of CD8 naïve T cells and T effector memory cells in hospitalized aged COVID-19 patients (**Fig2 b, d**), we examined a single-cell transcriptomic dataset of CD8 T cells^25^ in order to search for aging-related molecular mechanisms in a cell type-specific manner. Uniform Manifold Approximation and Projection^26^ (UMAP) revealed 5 distinct CD8 sub-clusters (**Fig. 3a, b**), including naïve CD8, T central memory (Tcm), Tem, CD8-proliferation, and CD8 T effector. Comparing to severe young COVID-19 patients, up-regulated genes (q < 0.05, log fold change > 0.1) in CD8 naïve T cells from aged patients formed a network module (the largest connected component) in the human protein-protein interactome (**Fig. 3c**). This age-specific network module was significantly enriched in several pathways, including apoptosis (q = 0.013), human T cell leukemia virus 1 infection (q = 0.013), and TNF signaling (q = 0.014, **Fig. 3c**). In particular, the apoptosis gene cathepsin D^27^ (*CTSD*) was highly expressed in naïve CD8 T cells from aged severe COVID-19 patients (q < 2.0 × 10^−16^). Down-regulated genes, such as interferon-stimulated genes *IFITM3* and *TRIM22*, in CD8 naïve T cells from aged COVID-19 patients were enriched in type I & II IFN signaling pathways (**Fig. 3c**). In addition, the transcription factor *STAT1*, an important downstream factor in type I & II IFN signaling pathways^28^, was down-regulated in CD8 naïve T cells in aged COVID-19 patients (**Fig. 3c**). Notably, the SARS-CoV-1 NSP1 protein impedes type I & II IFN signaling^29^ by attenuating STAT1 phosphorylation^30^. Thus, IFN deficiencies in CD8 naïve T cells may contribute to increased severity of COVID-19 disease in aged patients.

**Figure 3.**
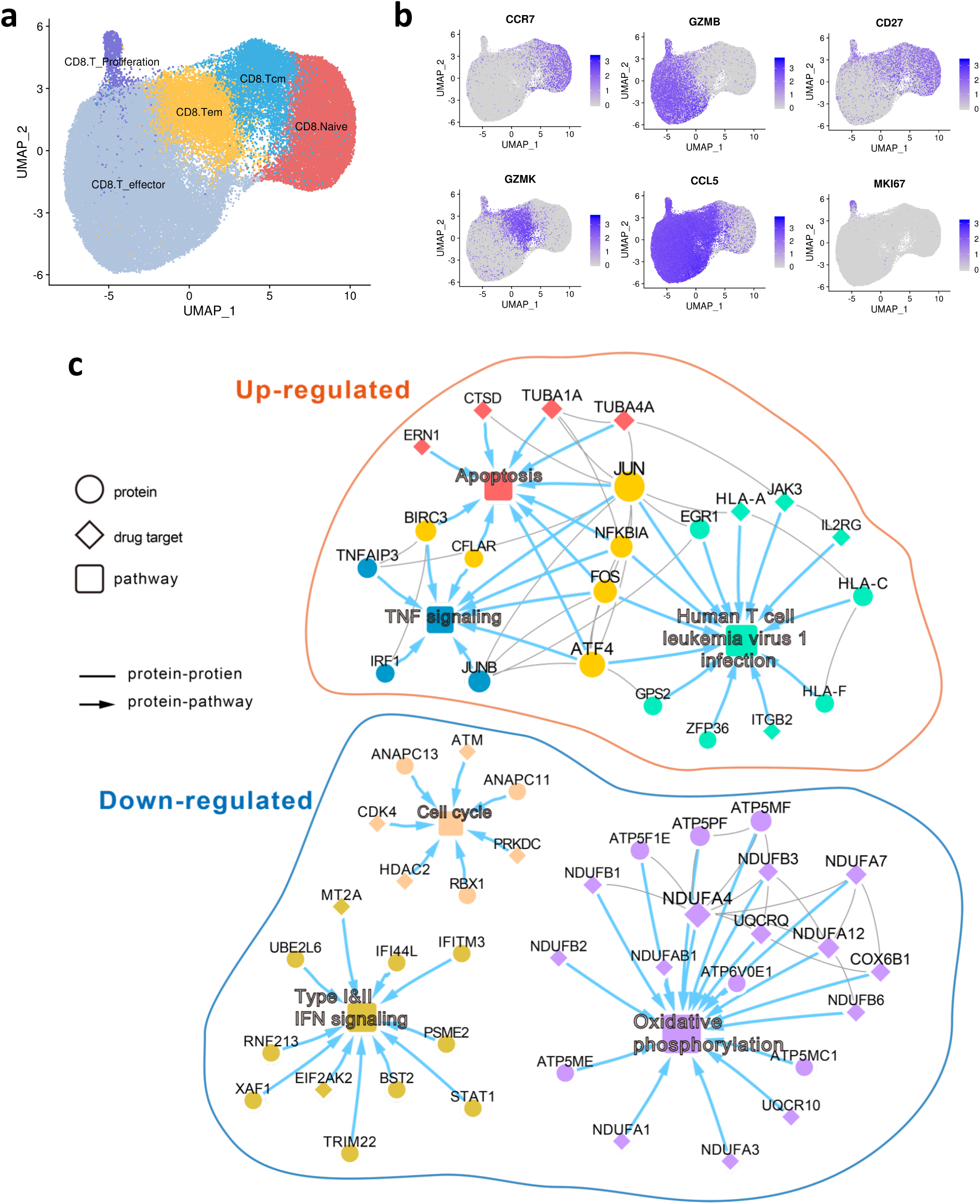
Single cell transcriptome of CD8 T cells in aged COVID-19 patients. **a** UMAP plot displays 8 identified CD8 T cell subpopulations. **b** The expression of marker genes shown on the UMAP plot. The expression levels are blue color coded. **c** A highlighted protein-protein interaction subnetwork for age-biased differentially expressed genes in CD8 naïve T cells from patients with critical COVID-19. The colors for nodes and edges represents different immune pathways.

### Interferon deficiencies correlate with SARS-CoV-2 viral load in aged patients

To further investigate the relationship between viral load and COVID-19 disease severity, we analyzed bulk RNA-seq data from nasopharyngeal samples^31^ (see Methods). Consistent with our findings in naïve CD8 T cells, expression levels of IFNα genes (*IFNA1, IFNA5, IFNA7* and *IFNA8*) were significantly decreased in aged patients with high viral load (**Fig. 4a**). In addition, expression of *IFNG* was decreased in aged patients with low viral load (**Supplementary Fig. 4**). Notably, we found that the IFN-stimulated antiviral genes,^32^ including *IFIT1* and *OAS1* (2’-5’-Oligoadenylate Synthetase 1), were down-regulated in aged patients with a higher viral load (**Fig. 4b**). Next, we performed gene set enrichment analysis (GSEA, see Methods) for differentially expressed genes in aged vs. younger individuals with a higher viral load and found downregulation of genes in the innate immune pathways (FDR < 0.05, **Fig. 4b**) of RIG-I like receptor signaling, Toll-like receptor signaling, and NOD-like receptor signaling in aged severe COVID-19 patients. RIG-I like receptors senses SARS-CoV-2 RNA and subsequently type-I IFN production^33^; however, SARS-CoV-2 has evolved several mechanisms to blunt IFN induction, including the direct targeting of MDA5 (melanoma differentiation-associated protein 5), a RIG-I-like receptor, by the viral papain-like protease (PLpro)^34^. Furthermore, IFN is potently inhibits IL-8 expression^35^ in viral infection, and we also showed that aged COVID-19 patients with high viral load exhibit elevated plasma IL-8 (p = 0.006, Mann–Whitney *U* test, **Fig. 4c**). Notably, up-regulated genes in aged patients with high viral load were not enriched in immune pathways (**Fig. 4b**), indicating decreased immune ability in response to SARS-CoV-2 infection. Taken together, our data show that IFN deficiency is associated with elevated SARS-CoV-2 viral load in aged patients.

**Figure 4.**
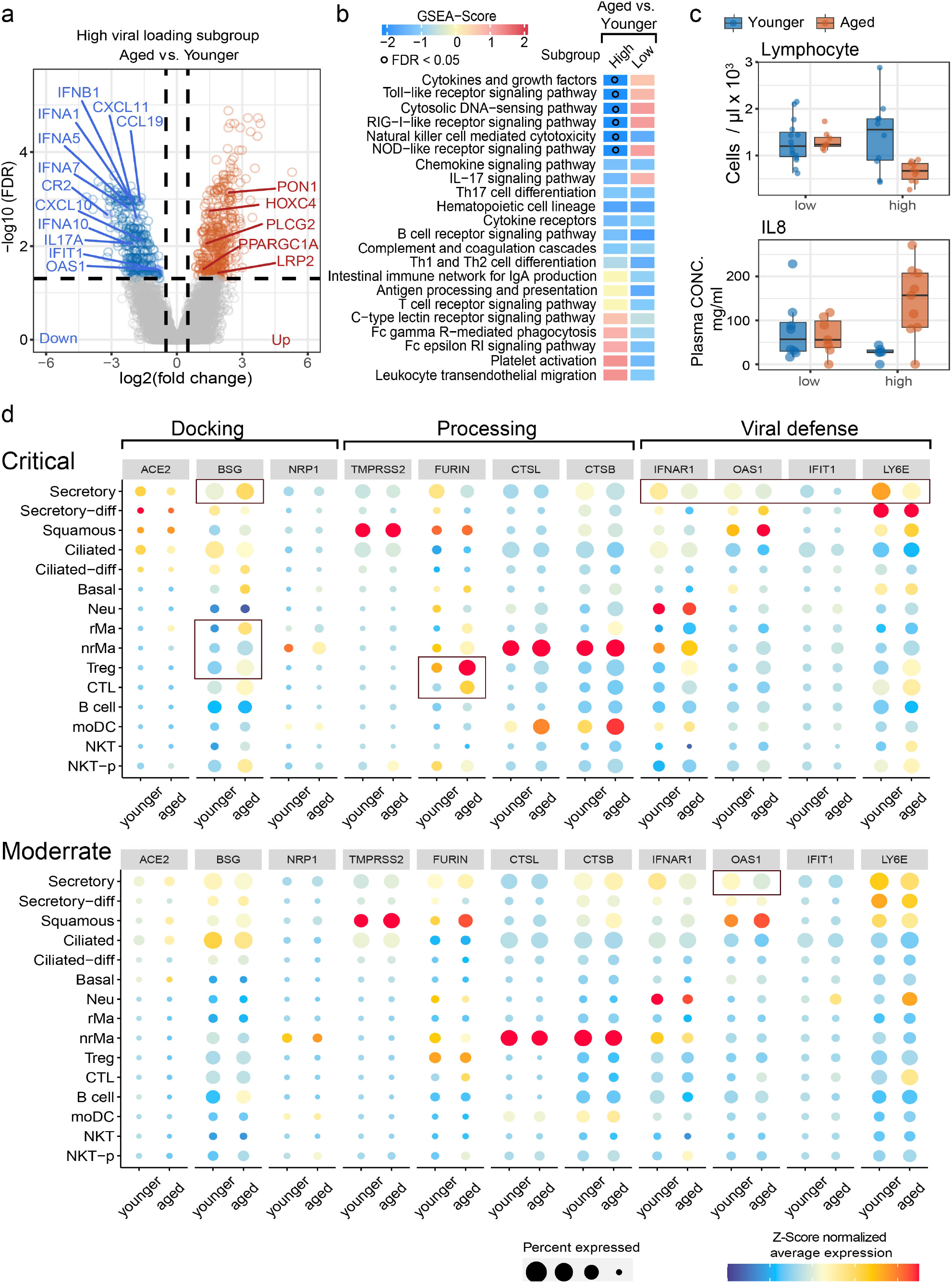
Analysis of SARS-CoV-2 viral load and related entry gene expression in nasal tissues. **a**, Volcano plot showing the differential genes of bulk RNA-sequencing data in aged versus younger patients in high viral load nasal tissues. **b**, Gene-set enrichment analysis (GSEA) of 22 immune pathways for differential gene set in two comparable conditions. The gradient color bar shows the NES score (see Method). NES score > 0 and q < 0.05 indicates that up-regulated DEGs in aged vs. young are significantly enriched in immune pathways, while NES score < 0 and q < 0.05 indicates down-regulated DEGs in aged vs. young are significantly enriched in immune pathways. Black dots denote FDR < 0.05. **c**, Boxplot showing the lab testing data changes in aged and younger COVID-19 patients with high (> 4.5 log10 RNA copies/ml) and low (< 4.5 log10 RNA copies/ml) viral load. **d**, SARS-CoV-2 related entry gene expression profile across 15 cell types of nasal tissue between aged and younger patients. The size of dot denotes the percentage of the positive cell which expressed the tested genes. The gradient color bar represents the z-score scaled average expression of genes in each cell type.

### Age-dependent increased expression of SARS-CoV-2 entry factors

We next investigated age- and cell type-specific expression of SARS-CoV-2 entry factors using a single-cell RNA-sequencing dataset^36^ (scRNA-seq, see Methods) from nasal tissue of critical (n = 11) and moderate COVID-19 patients (n = 8, see Methods). In total, the scRNA-seq dataset contained 115,895 cells across 15 well-annotated cell types within two main cell populations: epithelial cells (6 cell types) and immune cells (9 cell types).

We found that secretory and ciliated cells in aged COVID-19 patients display reduced abundance of angiotensin converting enzyme-2 (ACE2), a key SARS-CoV-2 docking receptor^37^ (**Fig 4d**). However, the more recently identified SARS-CoV-2 docking receptor basigin^38^ (BSG or CD147) was expressed in 95% of secretory cells in aged patients with critical COVID-19 (**Fig 4d**, and **Supplementary Table** 6), most notably in Treg (regulatory T cell) and CLT (cytotoxic T) cells (**Fig. 4d**). We also found that the S protein priming proteases TMPRSS2^39^ and FURIN^24^ were highly expressed in epithelial cells in critical and moderate COVID-19, with no differences between aged and young patients (**Fig 4d, Supplementary Table 6**). However, FURIN levels were increased in several immune cell types, including Treg and CLT, in aged patients with critical COVID19 (**Fig 4d**). Taken together, our results suggest that elevated expression of two SARS-CoV-2 factors (BSG and FURIN) in Treg and CLT cells may contribute to the increased susceptibility of aged patients to COVID-19.

### Increased immune-epithelial cell interactions in aged COVID-19 patients

To further investigate the immunological mechanisms underlying age-associated COVID-19 outcomes, we performed GSEA to explore transcriptomic signatures on 22 immune pathways across 15 cell types derived from nasal tissue (see Methods). Here, we observed distinct immune responses between older and younger individuals with critical or moderate COVID-19 (**Supplementary Fig. 6**) in epithelial and immune cell types. We next used CellphoneDB^40^ to quantify ligand-receptor interactions between epithelial and immune cells and found an elevated number of significant ligand-receptor interactions involved in immune-epithelial interactions (p < 0.05, permutation test, **Supplementary Table 7**) in aged patients with critical COVID-19 (**Fig. 5a)**. In addition, we also found a stronger immune-epithelial cell interaction network in aged patients. In particular, we noted that secretory-T regulatory cells (Treg) and non-resident macrophages (nrMa) displayed the highest connection with other cell types in aged patients with critical COVID-19 (**Fig. 5a**).

**Figure 5.**
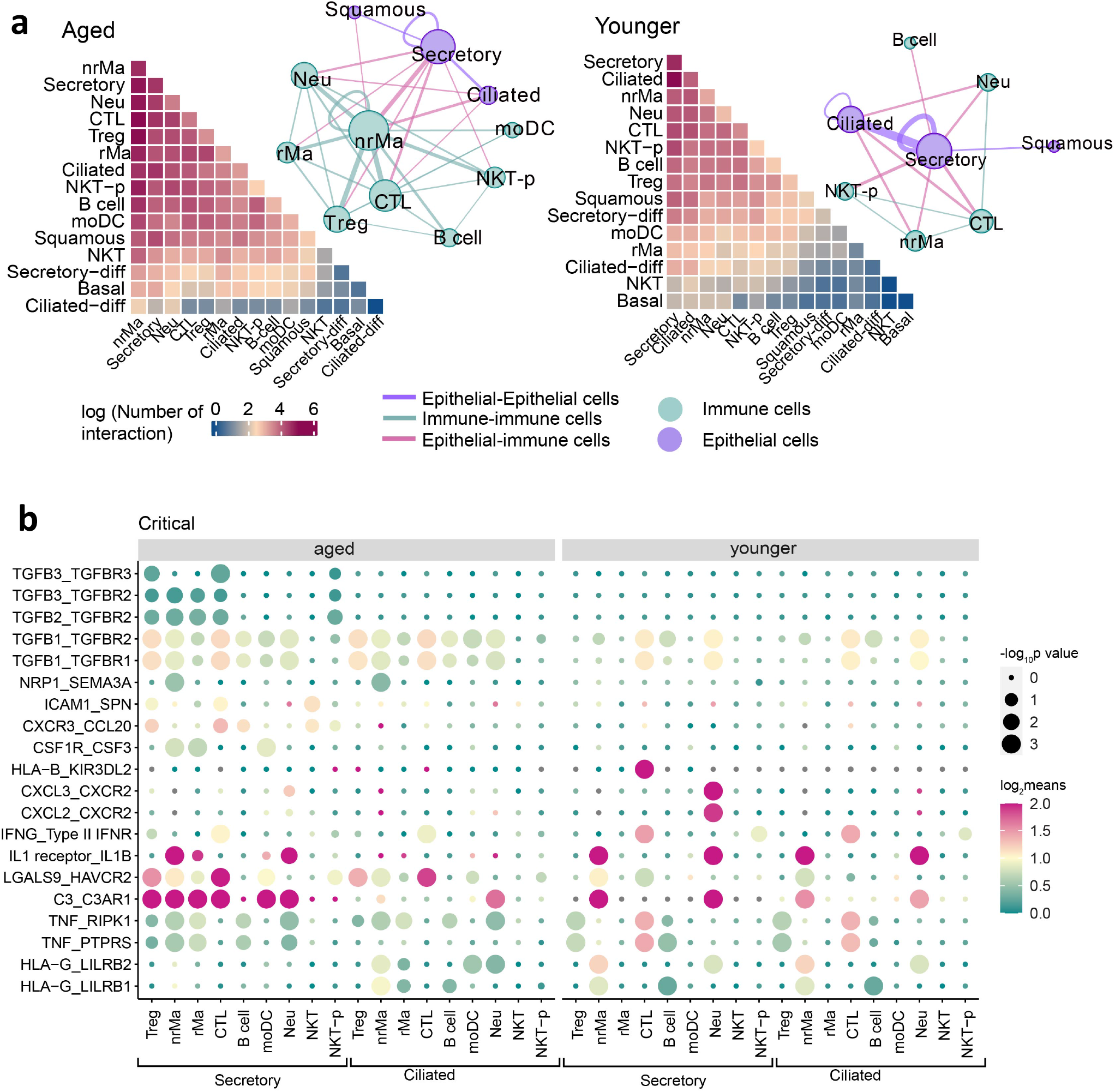
Distinct epithelial-immune cell interaction profile in aged and younger patients with critical COVID-19. **a**, Heatmap showing the total log-scaled interaction number between epithelial-immune cells in critical COVID-19 disease. Aged group, n= 3 patients, younger group, n = 5 patients. The cell-cell interaction network depicted all cell pairs in which the number of cell-cell interaction > 50. Edge size denotes the number of interactions between two cell types. Different colors indicate the immune or epithelial cell types. **b**, Dot plot showing significant ligand-receptor interactions between epithelial-immune cell interaction in critical COVID-19 disease. The circle size indicates -log10-scaled p values by permutation test, and gradient color bar shows the log2-scaled means of average expression of interacted cell pair.

We next analyzed ligand-receptor interactions of secretory/ciliated – immune cells in aged and younger patients with critical COVID-19 (**Fig. 5b**), and identified higher levels in aged patients (p < 0.05, permutation test). For example, we found elevated expression of TGF-β genes (*TGFB1, TGFB2* and *TGFB3*) and their interacting partners (i.e., *TGFBR2* and *TGFBR3*, p < 0.05, permutation test, **Supplementary Table 8**) in both Treg and nrMa cells. Of note, TGF-β has previously been shown to regulate the chronic immune response to SARS-CoV-2 in severe COVID-19 patients^41^. Thus, TGF-β-mediated strong secretory and Treg cell interaction may explain the longer duration of hospitalization in aged COVID-19 patients (**Supplementary Fig. 1b**).

We also observed distinct immune-epithelial cell interactions in younger COVID-19 patients. For example, secretory and CTL cells expressed high levels of several ligand-receptor pairs, including HLA-B – KIR3DL2, TNF – RIPK1 and TNF – PTPRS (p < 0.05, permutation test), and secretory and Neu cells highly co-expressed CXCL2/3 and CXCR2 (p < 0.05, permutation test). In addition, we found that secretory/ciliated – CTL cells showed a similar IFNG – IFNGR pattern, while the expression level in aged patients was much lower (**Fig. 5b**). In particular, secretory/ciliated – CTL cell interaction in younger patients showed strong IFNG – IFNGR interaction compared to aged patients with moderate COVID-19 (**Supplementary Fig. 6**). In summary, these observations revealed that immune-epithelial cell interactions are associated with critical COVID-19 in aged patients. In particular, reduced expression of IFNR signaling is associated with greater severity of COVID-19 in aged individuals (**Fig. 4a**).

## Discussion

This study provides a comprehensive analysis of immune profiles in aged and younger COVID-19 patients. Previous epidemiologic studies have identified age as an important risk factor for severe COVID-19^1,42,43^, and our large COVID-19 registry data further confirmed the elevated likelihood of severe COVID-19 in aged individuals even after adjusting for sex, race, smoking and multiple disease comorbidities (**Fig. 1c**). Mechanistically, aged hospitalized COVID-19 patients showed elevated levels of D-dimer, CRP (**Fig 1d**) and NLR (**Fig 1e**), which are inflammatory markers associated with severity and death in COVID-19^16,44^.

Via deep immune cell profiling data analysis, we identified distinct immune responses in younger and aged COVID-19 patients (**Fig. 6**). For example, both younger and aged COVID-19 patients showed increased ncMono cells and elevated IL6 (**Fig. 2, Fig. 6**), while only aged COVID-19 patients displayed elevated plasma IL8 and IL27 (**Fig. 2g** and **Supplementary Fig. 2b**). IL-6 is a potential therapeutic target since it is a critical mediator of cytokine storm in COVID-19^45^. However, a recent Phase III clinical trial (NCT04320615) showed no reduced mortality in severe COVID-19 patients treated with the anti-IL-6R monoclonal antibody tocilizumab^46^. Younger COVID-19 patients in the ICU also showed significantly higher IL10 (**Supplementary Fig. 2b**). Our observations suggest that targeting IL-10 might reduce mortality in younger patients with severe COVID-19, whereas IL-8- and IL-27-based therapies might benefit aged COVID-19 patients.

**Figure 6.**
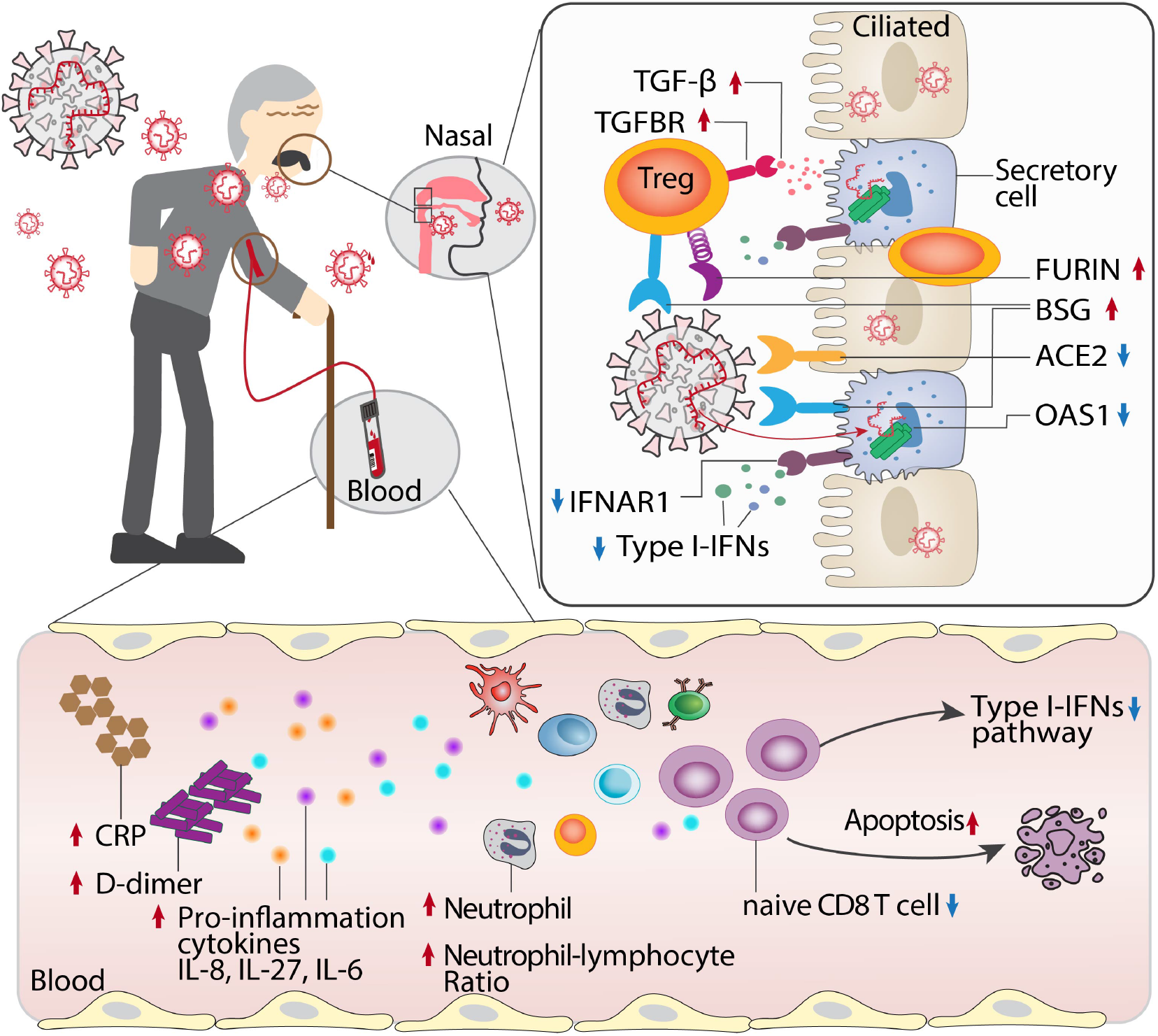
Proposed mechanistic models for age-biased COVID-19 severity in aged individuals. Several age-related pathophysiologic immune responses are associated with disease susceptibility and severity in COVID-19: a) decreased lymphocyte count and elevated inflammatory markers (C-reactive protein [CRP], D-dimer, and neutrophil-lymphocyte ratio); b) elevated pro-inflammation cytokines IL-8, IL-27 and IL-6 in aged COVID19 patients; c) reduced abundance of naïve CD8 T cells with decreased expression of antiviral defense genes (i.e., *IFITM3* and *TRIM22*) in aged individuals with severe COVID-19; d) type I interferon deficiency is associated with SARS-CoV-2 viral load in aged individuals; e) elevated expression of SARS-CoV-2 entry factors (*BSG* and *FURIN*) and reduced expression of antiviral defense genes (*IFNAR1, OAS1, IFIT1*) in the secretory cells of critical COVID-19 in aged individuals; f) strong TGF-beta mediated immune-epithelial cell interactions (i.e., secretory – T regulatory cells) in aged individuals with critical COVID-19.

We also found reduced lymphocytes in hospitalized aged COVID-19 patients (**Fig. 1d**). In particular, the abundance of naïve CD8 T cells was decreased in aged patients with severe COVID-19 (**Fig. 2d**). Reduction of naïve CD8 T cell is a hallmark of immunosenescence in older individuals^47^, and through scRNA-seq data analysis we observed significant enrichment of upregulated apoptosis genes in CD8 naïve T cells from aged COVID-19 patients. Mechanistically, the apoptosis driver gene *CTSD*^*27*^ is significantly elevated in naïve CD8 T cells from aged severe/critical COVID-19 patients compared to younger patients (q < 2.0 × 10^−16^). Thus, modulation of CD8 naïve T cell dysfunction, especially targeting apoptosis pathway^48^, may provide a new treatment strategy for severe COVID-19 in aged patients.

IFN-mediated immunity provides initial rapid protection against viral infection^49^, and about 3.5% of patients with life threatening COVID-19 show genetic aberrations in the type I IFN pathway^50^. A recent genetic study in European ancestry revealed that the cis-protein quantitative trait loci (pQTL, rs4767027) in *OAS1* (an IFN-stimulated gene) was significantly associated with decreased likelihood of COVID-19 susceptibility and severity^51^. Herein, we found that aged individuals with severe COVID-19 show reduced expression of type I IFN genes (**Fig. 3c, 4a, 5b**). Notably, aged patients with high SARS-CoV-2 viral load show reduced expression of *OAS1* and *IFNA1, IFNA5, and IFNA7* (**Fig. 4a**) compared to younger patients. On the contrary, aged patients with high SARS-CoV-2 viral load have elevated expression of the pro-inflammatory cytokine IL-8 and decreased lymphocyte cell counts in plasma (**Fig. 4c**), demonstrating dysregulation of cytokine responses that has been well described for COVID-19^52^. Of note, the dysregulated cytokine response is likely the effect of a variety of immunomodulatory strategies employed by SARS-CoV-2 that are used to manipulate specific signaling pathways that lead to cytokine induction such as the RIG-I-like receptor pathway. Therefore, the type I IFN pathway offers a potential therapeutic target for aged severe COVID-19 patients.

Although aged adults show increased susceptibility to SARS-CoV-2 infection compared to children^5^, we did not find differences in SARS-CoV-2 viral load in the upper airways between younger and aged patients (**Supplementary Fig. 7**). Using large-scale scRNA-seq data analysis, we did find, however, that the SARS-CoV-2 entry genes (*ACE2, BSG, TMPRSS2, FURIN* and *NPR1*) showed cell type-specific expression profiles in both aged and younger individuals. In aged patients with critical COVID-19, expression of BSG was increased in secretory, rMa and CTL cells, and elevated expression of FURIN was found in Treg and CTL cells. Thus, cell type-specific host factor expression may play an important role in age-mediated disease susceptibility and severity in COVID-19.

We also identified age-specific increases in immune-epithelial cell interactions. For example, we found strong TGF-β mediated immune-epithelial cell interactions in aged severe COVID-19 patients. TGF-β plays a crucial role in pulmonary fibrosis^53,54^, which is a common complication in severe COVID-19 patients^55^. The nucleocapsid protein of SARS-CoV-1 also upregulates TGF-β expression^56^. Thus, TGF-β targeted therapies may be of utility in aged patients with COVID-19. We additionally identified receptor interacting serine/threonine kinase 1 (RIPK1)-mediated immune-epithelial cell interactions (secretory/ciliated -CTL cell pairs) in younger patients with critical COVID-19. RIPK1 is a key mediator of inflammation^57^, and a RIPK1 inhibitor (SAR443122) has been tested in a phase I clinical trial (ClinicalTrials.gov Identifier: NCT04469621) to treat tissue damage resulting from inflammation in severe COVID-19 patients. Altogether, RIPK1 inhibitors^58^ may offer a potential treatment for young COVID-19 patients, such as COVID-19 related multisystem inflammatory syndrome in Children (MIS-C) in children^59^.

Lastly, we acknowledge the potential limitations of our study. Although we inspected omics data from multiple tissues, including PBMCs, plasma, and nasal tissues, additional analysis of other COVID-19 and aging relevant tissues, such as lung and brain, should be investigated in the future. In addition, our COVID-19 database and omics data were generated from acute COVID-19 patients, and identification of the underlying genetic and molecular basis of aging differences for long-haul COVID-19 patients will be an important area of future investigation^60^. Finally, investigation of COVID-19 vaccine responses between aged and young patients are also warranted in the future.

## Methods and Materials

‘Young’ was defined as 18 to 55 years of age, and ‘aged’ was defined as ≥ 65 years old.

### U.S. CDC COVID-19 epidemiological data

Publically-accessible COVID-19 death counts in 54 states in United States (U.S.) were downloaded from the Centers for Disease Control and Prevention (CDC) website (https://data.cdc.gov/NCHS/Provisional-COVID-19-Death-Counts-by-Sex-Age-and-S/9bhg-hcku/data) on December 23th, 2020 (**Supplementary Table 1**). Publically-accessible statistics of influenza mortality across 10 flu seasons (November, 2010 - 2020) in U.S. was downloaded from CDC website (https://catalog.data.gov/dataset/deaths-from-pneumonia-and-influenza-pi-and-all-deaths-by-state-and-region-national-center-) on June 20^th^, 2020. Both COVID-19 and influenza datasets include three age-stratified groups: 0-17 years, 18-64 years, 65 years and older. These datasets were used for epidemiological prevalence analysis of COVID-19 and influenza.

We collated U.S. COVID-19 Case Surveillance Public Use Data from the CDC website (https://healthdata.gov/dataset/covid-19-case-surveillance-public-use-data) from December, 2019 to December 28^th^, 2020. This dataset includes age-stratified COVID-19 case counts in hospitalization, ICU admission, death, sex, and race. We extracted two age subgroups from all laboratory-confirmed cases using the following criteria: i) the age range of younger group from 20 to 49 years, and the age range of older group over than 60 years (**Supplementary Table 2**); ii) deletion of all cases in which sex and race information was missing. In total, the younger subgroup includes 2,369,919 cases, with 94,161 in hospitalization, 9,138 in ICU admission, and 6,469 death cases. The older subgroup has 1,048,011 cases in total, with 243,109 in hospitalization, 29,671 in ICU admission, and 124,566 death cases. This dataset was used to determine odds ratio analysis.

### COVID-19 registry

We used institutional review board–approved COVID-19 registry data, including 45,077 individuals (12,651 were aged patients and 32,426 were younger patients, Supplementary Table 3) tested during March to December, 2020 from the Cleveland Clinic Health System in Ohio and Florida. All tested samples were pooled nasopharyngeal and oropharyngeal swab specimens. Infection with SARS-CoV-2 was confirmed by RT-PCR in the Cleveland Clinic Robert J. Tomsich Pathology and Laboratory Medicine Institute. In total, 12,304 patients (aged n= 3,559, younger n = 8,745) tested COVID-19 positive by the end of December, 2020. All SARS-CoV-2 testing was authorized by the Food and Drug Administration under an Emergency Use Authorization, in accord with the guidelines established by the Centers for Disease Control and Prevention.

The data in COVID-19 registry includes COVID-19 test results, baseline demographic information, and all recorded disease conditions (Supplementary Table 3). We conducted a series of retrospective studies to test the association of aging with COVID-19 outcomes, including hospitalization, ICU admission, mechanical ventilation, and death. Data were extracted from electronic health records (EPIC Systems), and patient data was managed using REDCap electronic data capture tools. To ensure data quality, a study team trained on uniform sources for the study variables manually checked all datasets. Statistical analysis for smoking, hypertension, diabetes, coronary artery disease asthma, and emphysema & chronic obstructive pulmonary disease [COPD] were calculated after missing value deletion.

### Clinical outcome analysis

The odds ratio (OR) was used to measure the association between COVID-19 outcomes and aging based on logistic regression. An OR > 1 indicates that aged patients are associated with a higher likelihood of the outcome. To reduce the bias from confounding factors, we employed OR analysis in two datasets. For U.S. CDC datasets, the OR model was adjusted by sex and race, due to limited information of other confounding factors. However, in the COVID-19 registry we adjusted for sex, race, smoking, hypertension, diabetes, coronary artery disease, asthma, emphysema, and chronic obstructive pulmonary disease. The Kaplan-Meier method was used to estimate the cumulative hazard of hospitalization of COVID-19 patients across age groups. For hospitalization outcome, the time was calculated from the start date of COVID-19 symptoms to hospital admission date. Log-rank test was used for comparison across different age groups with Benjamini & Hochberg adjustment. Cumulative hazard analysis was performed using the Survival and Survminer packages in R 3.6.0 (https://www.r-project.org).

### Published available COVID-19 datasets were used in this study

Detailed information of the list datasets id shown in **Supplementary Table 4**.

#### Two single-cell sequencing datasets

In this study, we used two COVID-19 single-cell datasets (**Supplementary Table 1**). First, the CD8^+^ T cells dataset^25^ is a sub-dataset from original PBMC single cell data^61^. We re-analyzed 59,815 single cell transcriptomes of CD8 T cells, which revealed 5 distinct CD8 sub-clusters (**Fig. 3a**), including CD8 naïve (CCR7^+^, LEF1^+^), Tcm (GZMK^+^,LTB^+^, CCR7^-^), Tem (GZMK^+^, CCR7^-^), CD8-proliferation (MKI67^+^), CD8 T effector (ZNF683^+^, GZMB^+^, GNLY^+^). Based on our aging criteria, the final subpopulation included 32 aged patients (mild/moderate, n = 20; critical/severe, n = 12) and 40 younger patients (mild/moderate, n = 27; critical/severe, n = 13). Second, a single cell dataset from nasal tissues^36^ (European Genome-phenome Archive repository: EGAS00001004481) was from COVID-19 positive patients (11 critically ill patients and 8 moderately ill patients). Based on our aging criteria, we extracted a subpopulation from the original cohort. The final COVID-19 cohort used in this study included 8 critically ill patients (5 younger and 3 older patients) and 7 moderately ill patients (4 younger and 3 older patients). As the original dataset supplied cell type information, additional analysis was based on cell type annotation. The dataset contained 115,895 cells across 15 cell types (B cell, Basal, Ciliated, Ciliated-diff, CTL, moDC, Neu, NKT, NKT-p, nrMa, rMa, Secretory, Secretory-diff, Squamous and Treg).

#### Bulk RNA-sequencing dataset in nasal tissue^31^

The dataset was publically available from NCBI GEO database (GSE152075). Based on original meta-information, we extracted COVID-19 positive sample data with high or low viral load, deleting samples in which sex and age information were missing. 147 bulk RNA-seq samples were used in this study, including 61 aged patients (high viral load n = 27, low viral load n =34) and 86 younger patients (high viral load n = 46, low viral load n =40).

#### SARS-CoV-2 viral load dataset^62^

We quantified SARS-CoV-2 RNA load from 5 specimen types, including upper airway specimens (oropharyngeal swab [detectable percentage was 67%], nasopharyngeal [detectable percentage was 50%], sputum [detectable percentage was 85%]), plasma (detectable percentage was 27%), and urine (detectable percentage was 10%). We selected hospitalized patients with at least one COVID19-positive test among upper airway or plasma specimens. Finally, 72 patients were used for correlation analysis between age and viral loading. 43 patients (older patients n = 18, younger patients n = 25) with SARS-CoV-2 RNA detectable testing in upper airway were used to analyze the change of clinical inflammatory variables in both aged and younger groups. In our study, 54 patients tested positive for plasma SARS-CoV-2 RNA, including 21 patients with SARS-CoV-2 RNA (aged patients n = 13). There were 35 SARS-CoV-2 RNA undetectable patients (aged patients n = 7).

#### Circulating cell flow cytometry datasets^17^

This dataset included 12 major immune cell types as a percentage of PBMC and 32 T cell subtypes as a percentage of CD3 positive cells through flow cytometry (Supplementary Table 8). It also detected the plasma concentration of 71 cytokines through cytokine array. Based on our age criteria, the dataset included 81 hospitalized patients, 40 with longitudinal data. When the second follow-up time of a patient was greater than 7 days, it was recorded as two samples. Hence, 114 samples were analyzed, which included 94 older samples (non-ICU n = 66, ICU = 26) and 50 younger samples (non-ICU n = 37, ICU = 13).

### Single-cell sequencing data analyses

All single-cell data analyses and visualizations were performed with the R package Seurat v3.1.4 40. The data quality filtering was strictly followed by the original literature^19,36^. “NormalizeData” was used to normalize the data.

“FindIntegrationAnchors” and “IntegrateData” functions were used to integrate cells from different samples. Principal component analysis (PCA) and t-distributed Stochastic Neighbor Embedding (tSNE) with 15 principal components were used. A resolution of 0.5 was used in ‘FindClusters()’ step. ‘FindAllMarkers’ function with the MAST test was employed as the finding maker method for each cell type.

### Cell-cell interaction analysis

Cell-cell interactions analysis was based on normalized expression data of known ligand–receptor pairs in 15 cell types of nasal single cell sample. The analysis was performed by CellPhoneDB^40^ v2.1.4 (https://github.com/Teichlab/cellphonedb) based on the python 3.7 platform. Statistical analysis mode was used to identify significant ligand–receptor pairs in each cell number. A permutation test with 1,000 times were used to evaluate the significance.

### Bulk RNA-sequencing data analysis

All bulk RNA-sequencing data analysis started from raw counts value. R package edgeR^63^ v3.12 was used to analyze differentially expressed genes in older vs. younger groups. Correction for sex and batch effects was added into the formula of design model. Statistical significance p values were adjusted by Benjamini–Hochberg (q value) method. Differentially expressed genes were identified as adjusted p value (q) < 0.05 and log fold change > 0.5.

### Immune gene set enrichment analysis

To evaluated the immune pathway profiles in young and aged COVID-19 patients, GSEA was conducted as previously described^64^. Immune gene profiles were retrieved from the KEGG database^65^. We selected 22 immune-related pathways and 1241 genes from KEGG belonging to the immune system subtype. For each cell type, we performed a GSEA on the list of differential expressed genes (DEGs) ranked by the logr_2_FC. The normalized enrichment score (NES, Equation 1) was calculated for 22 immune pathways in young and aged specific gene sets (**Supplementary Fig 2a**),w

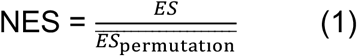

in which ES^64^ denotes enrichment score. Normalization of the enrichment score reduced the effect of the differences in gene set size and in correlations between gene sets and the expression dataset. NES score > 0 and q < 0.05 indicates that up-regulated DEGs in aged vs. young are significantly enriched in immune pathways, while NES score < 0 and FDR < 0.05 indicates down-regulated DEGs in aged vs. young are significantly enriched in immune pathways. Permutation test (1000 times) was performed to evaluate the significance. All analyses were performed with the prerank function in GSEApy package (https://gseapy.readthedocs.io/en/master/index.html) on Python 3.7 platform.

### Statistical analysis

Statistical tests for assessing categorical data through χ2 test, and the two-tailed Mann– Whitney *U* test was used to compare the difference of continuous variable by aged vs. younger. Spearman’s ρ was assessed for correlation between two variables. Statistical significance level was set at p < 0.05 and corrected by Benjamini–Hochberg (false discovery rate) method. All statistical analysis was performed by SciPy Statistics (https://docs.scipy.org/doc/scipy/reference/stats.html#module-scipy.stats).

## Data Availability

The clinical and transcriptomic datasets used in this study are public available, the details see Supplementary Table 1. The code for single cell analysis can be found in https://github.com/ChengF-Lab/COVID-19_Map.

## Author contributions

F.C. conceived the study. Y.H. performed all data processing and analyses. Y.Z., M.U.G., Y.L., L.J., T.C., H.Y., C.N., and A.A.P. discussed and interpreted all results. Y.H. and F.C. wrote and all authors critically revised the manuscript and gave final approval.

## Availability of data and materials

The clinical and transcriptomic datasets used in this study are public available, the details see **Supplementary Table 1**. The code for single cell analysis can be found in https://github.com/ChengF-Lab/COVID-19_Map.

---

**Supplementary Figure 1.**
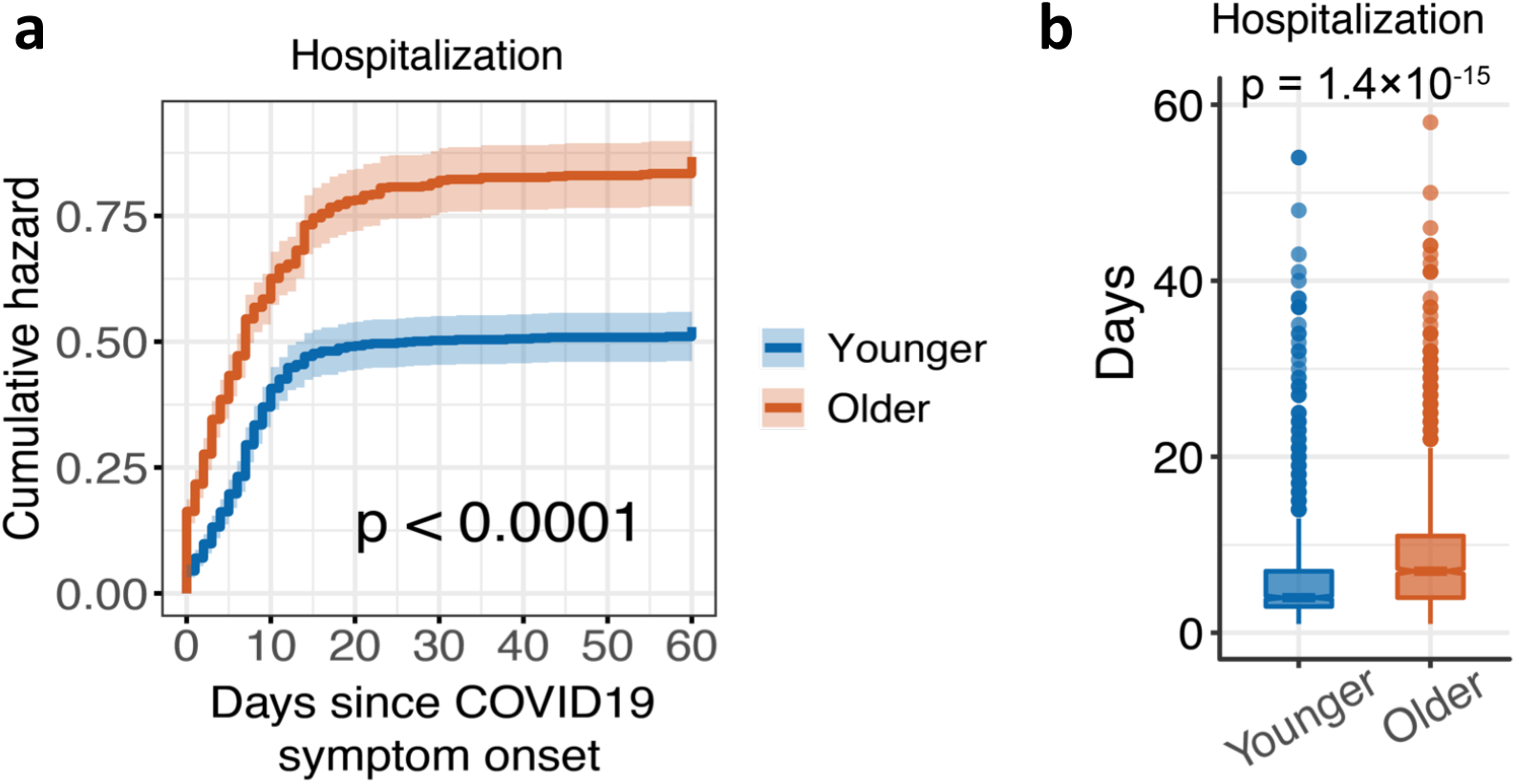
Aged COVID-19 patients with elevated hazard of Hospitalization. **a**, Cumulative hazard of COVID-19 hospitalization. The log-rank test with the Benjamini & Hochberg (BH) adjustment are used to compare the statistical significance of cumulative hazard of hospitalization. The shadow represents 95% confidence interval. **b**, Boxplots of the straying duration in hospital between aged (older) and younger patients. Statistical p-value was computed by Mann–Whitney *U* test.

**Supplementary Figure 2.**
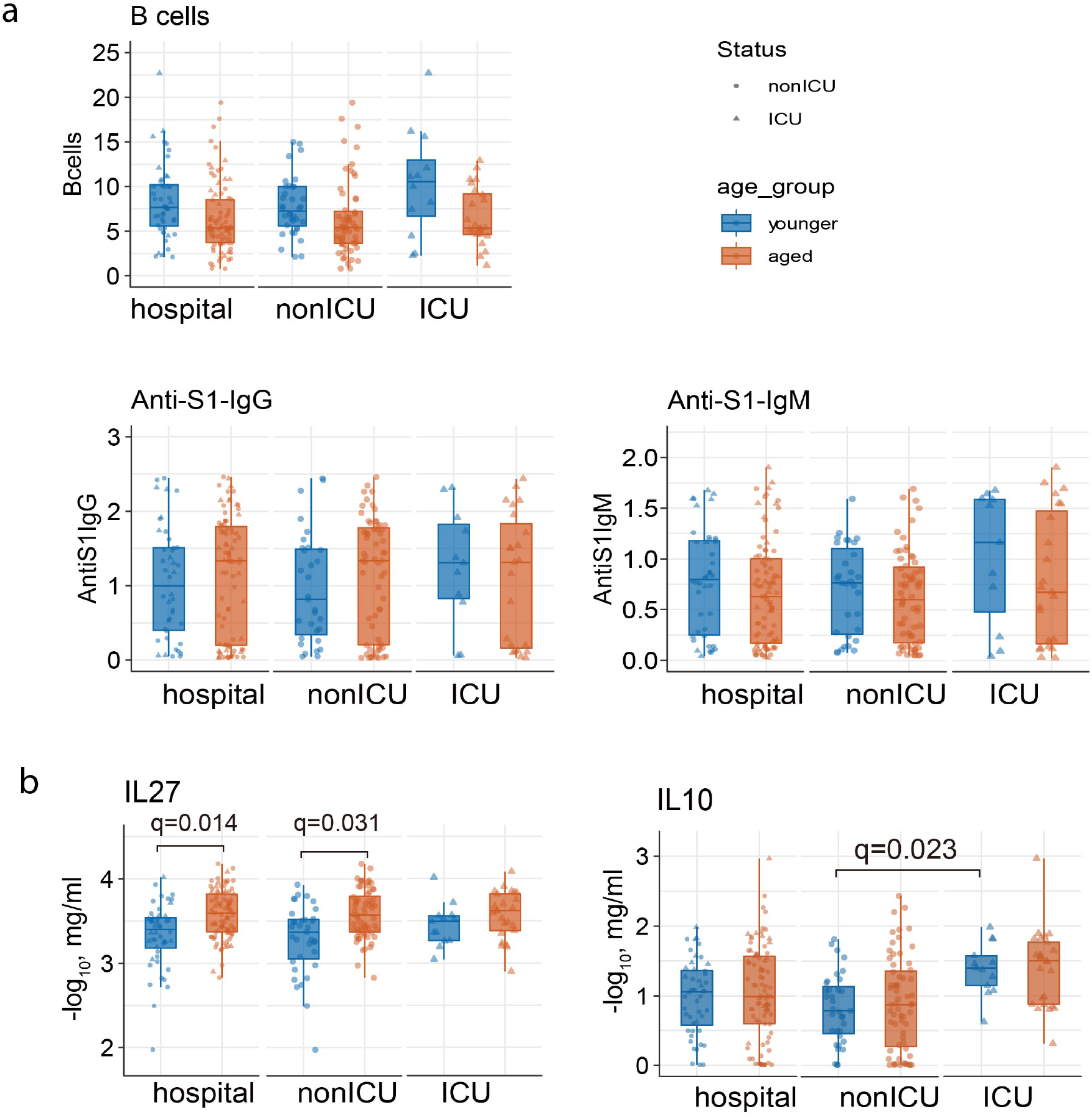
Comparison of the abundance of B cells in PBMCs and cytokines in plasma between aged and younger COVID-19 patients. **a**, The abundance of B cells subtype in all CD3 positive cells. **b**, Boxplot showing the plasma levels of cytokines and chemokines between younger and aged COVID-19 patients. Statistical p-value was computed by Mann–Whitney *U* test.

**Supplementary Figure 3.**
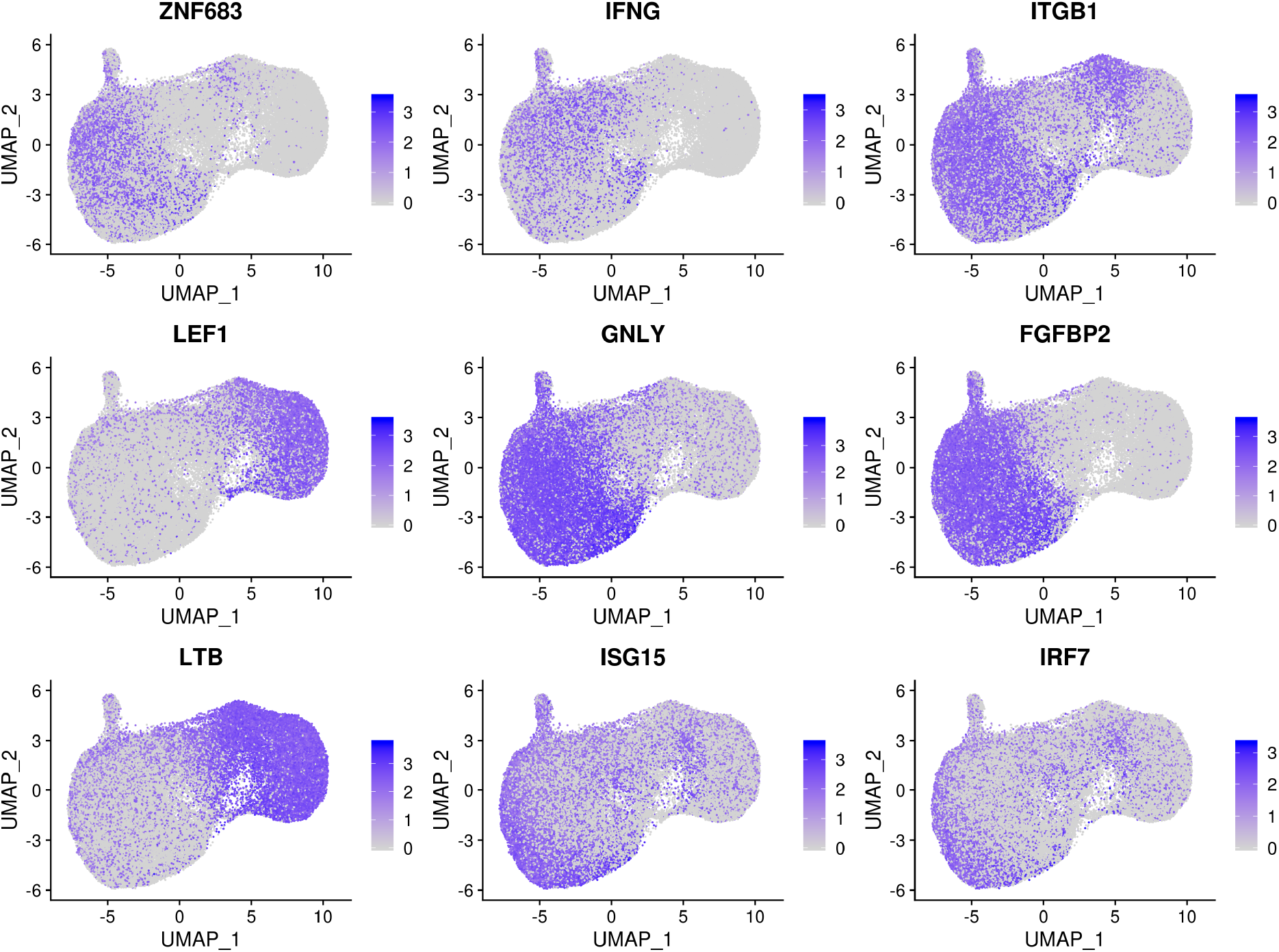
The expression of marker genes shown on the UMAP plot. The expression levels are blue color coded.

**Supplementary Figure 4.**
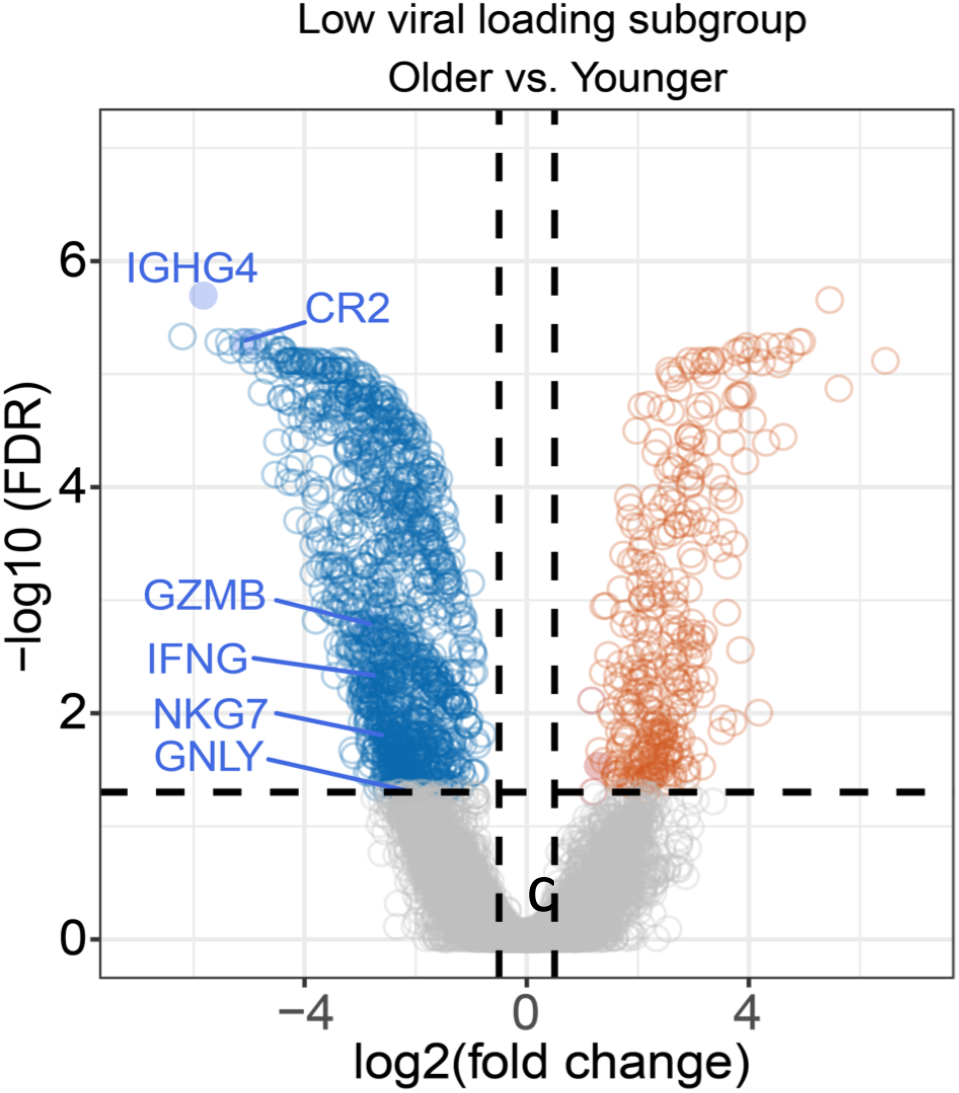
Analysis of relationship between age and SARS-CoV-2 viral load in nasal tissues. Volcano plot show the differential genes of bulk RNA-sequencing data in aged (older) versus younger in low viral load nasal tissues.

**Supplementary Figure 5.**
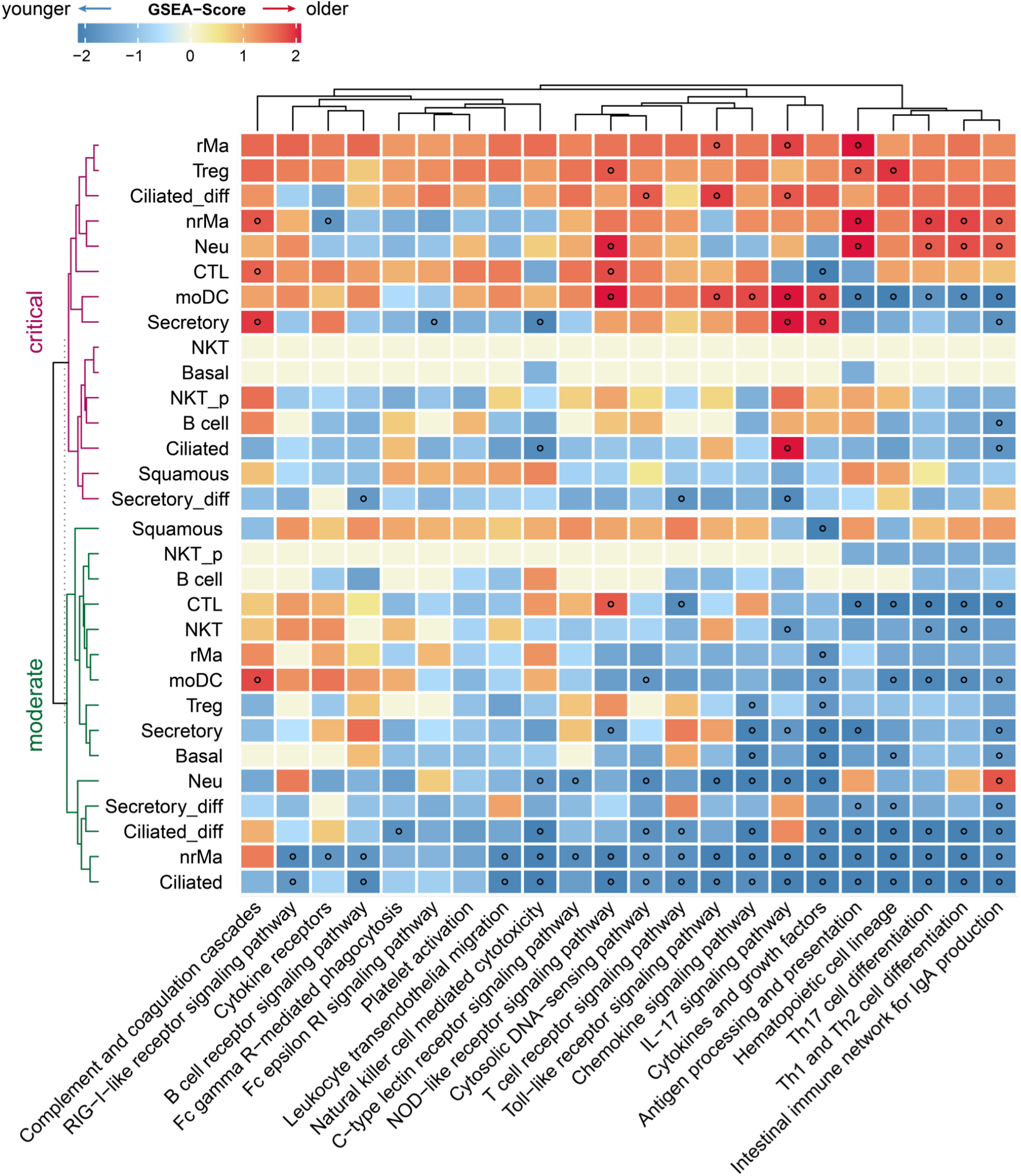
Gene-set enrichment analysis (GSEA) of 22 immune pathways across 15 cell types of nasal tissues. The gradient color bar shows the normalized enrichment score (NES) score. NES score > 0 indicates the immune pathway significantly enriched in upper-regulated genes. NES score < 0 indicates the immune pathway significantly enriched in down-regulated genes. Black dots denote the FDR < 0.05.

**Supplementary Figure 6.**
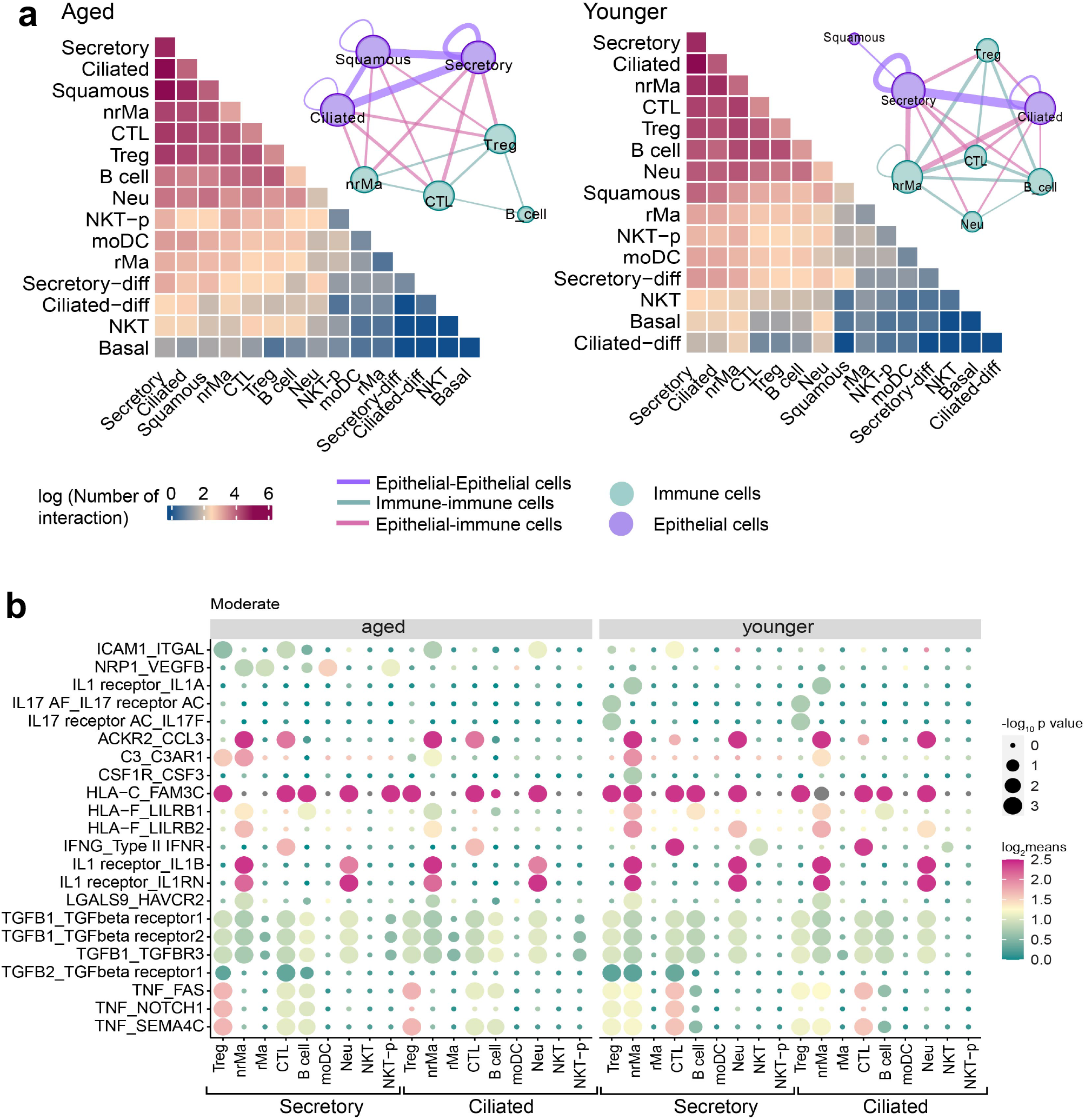
Distinct epithelial-immune cell interaction profile in aged and younger patients with moderate COVID-19. **a**, Heatmap show the total log-scaled interaction number between epithelial-immune cells in moderate COVID-19 disease. Aged group, n= 3 patients, younger group, n = 4 patients. The cell-cell interaction network depicted all cell pairs which the number of cell-cell interaction > 50. Edge size denotes the number of interactions between two cell types. Different colors indicate the immune or epithelial cell types. **b**, Dot plot showing the significant ligand-receptor interactions between epithelial-immune cell interaction in moderate COVID-19 disease. The circle size indicated -log10-scaled p values by permutation test. Gradient color bar shows the log2-scaled means of average expression of interacted cell pair.

**Supplementary Figure 7.**
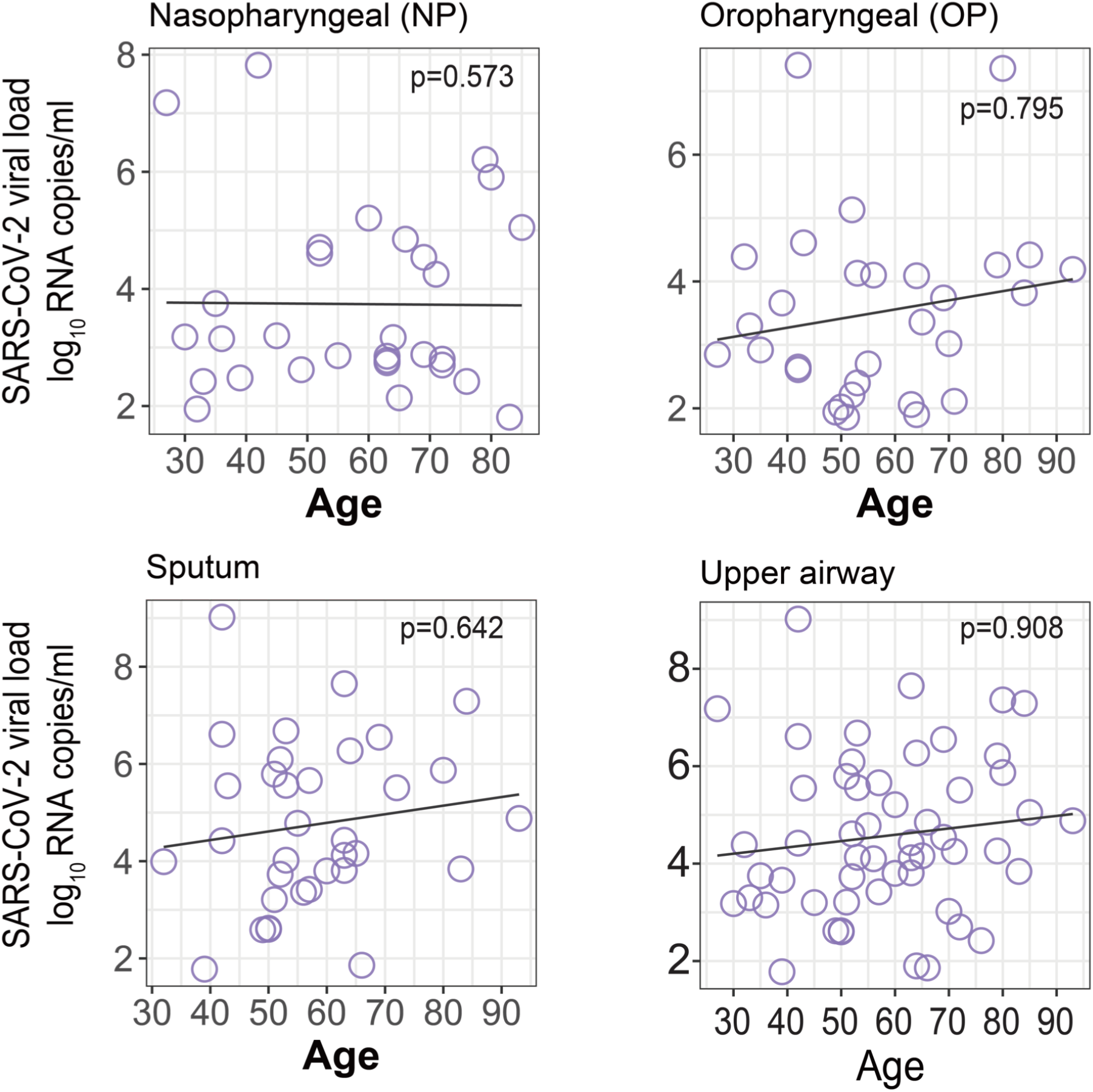
Correlation analysis between age and upper airway viral load. The upper airway data from three sample source, oropharyngeal swab, nasopharyngeal and sputum.

