## Supplementary_Figures1-7 for "Aging-related cell type-specific pathophysiologic immune responses that exacerbate disease severity in aged COVID-19 patients"

#### Supplementary Figure 1

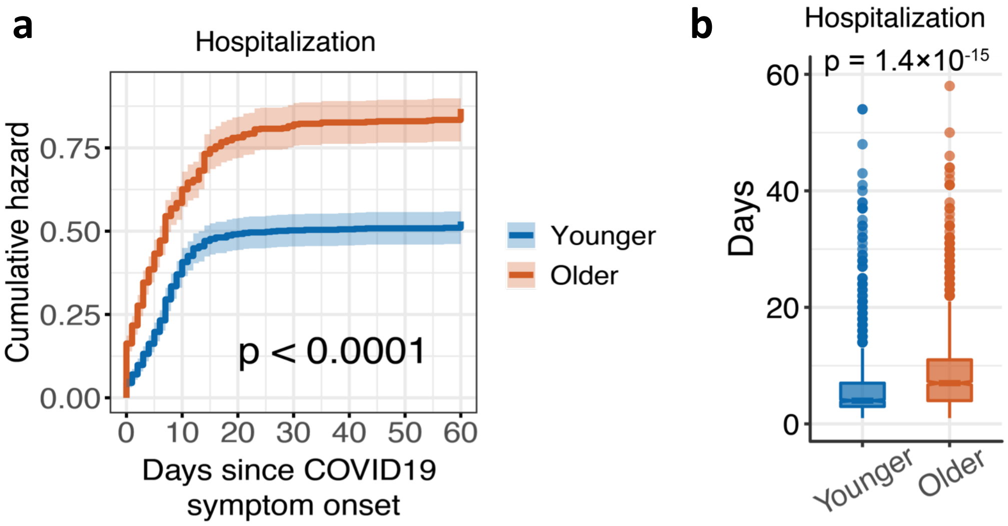


**Supplementary Figure 1. Aged COVID-19 patients with elevated hazard of Hospitalization. a,** Cumulative hazard of COVID-19 hospitalization. The log-rank test with the Benjamini & Hochberg (BH) adjustment are used to compare the statistical significance of cumulative hazard of hospitalization. The shadow represents 95% confidence interval. **b,** Boxplots of the straying duration in hospital between aged (older) and younger patients. Statistical p-value was computed by Mann–Whitney *U* test.

### Supplementary Figure 2

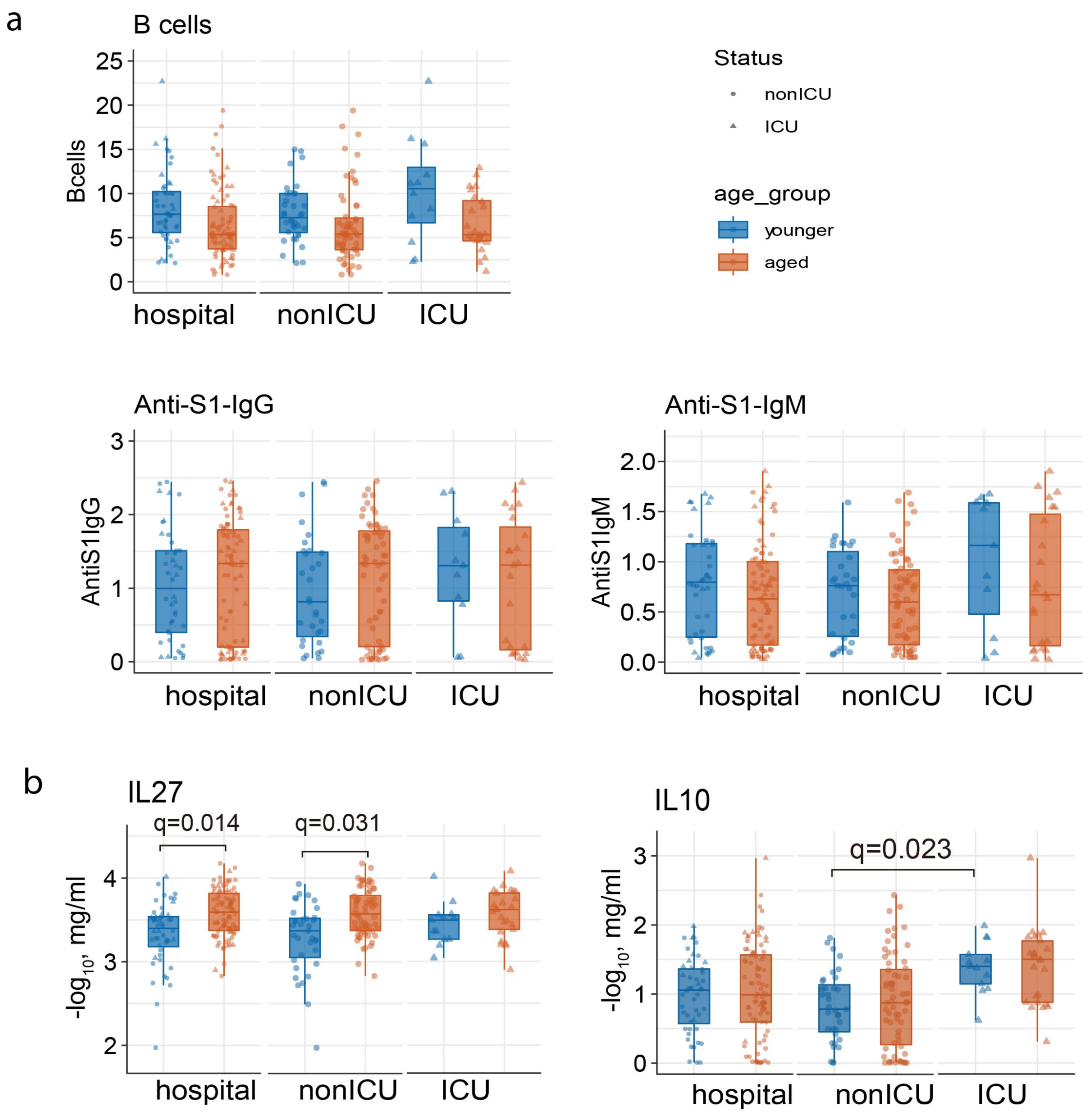


**Supplementary Figure 2. Comparison of the abundance of B cells in PBMCs and cytokines in plasma between aged and younger COVID-19 patients. a,** The abundance of B cells subtype in all CD3 positive cells. **b,** Boxplot showing the plasma levels of cytokines and chemokines between younger and aged COVID-19 patients. Statistical p-value was computed by Mann–Whitney *U* test.

### Supplementary Figure 3

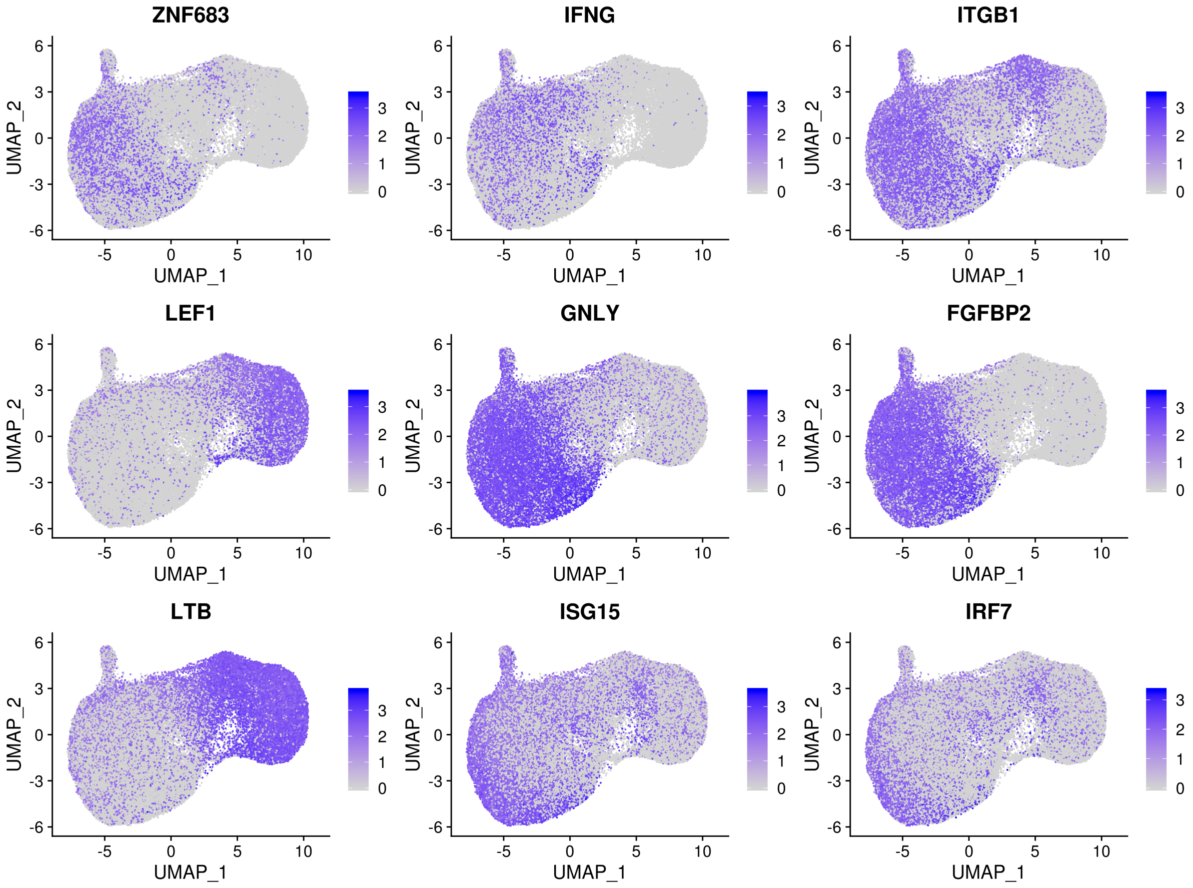


**Supplementary Figure 3. The expression of marker genes shown on the UMAP plot.** The expression levels are blue color coded.

### Supplementary Figure 4

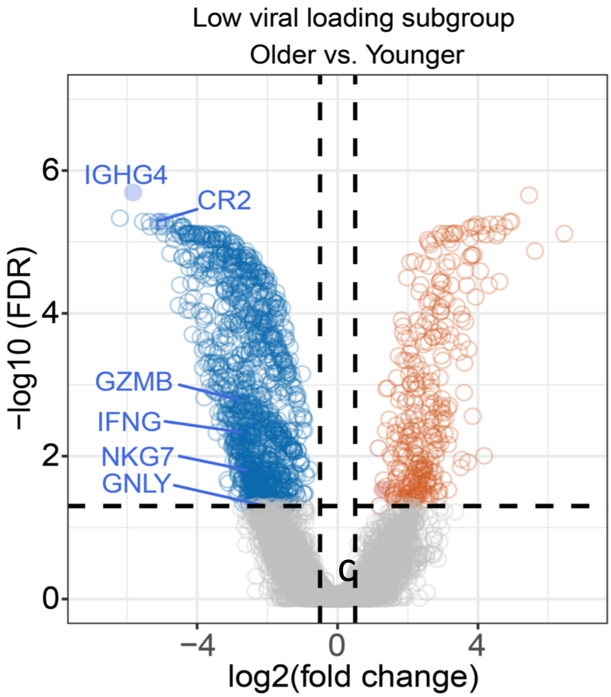


**Supplementary Figure 4. Analysis of relationship between age and SARS-CoV-2 viral load in nasal tissues.** Volcano plot show the differential genes of bulk RNA-sequencing data in aged (older) versus younger in low viral load nasal tissues.

### Supplementary Figure 5

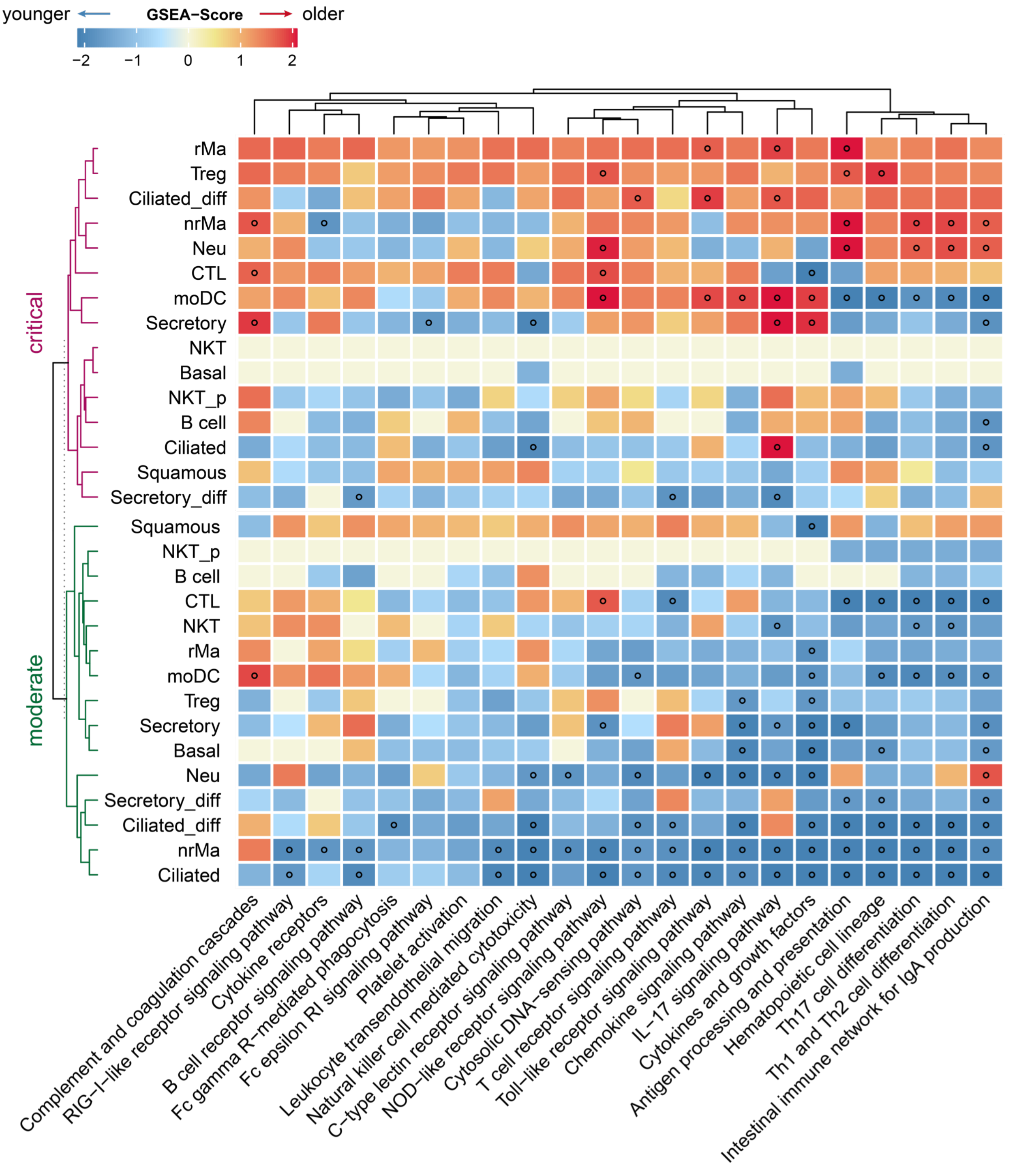


**Supplementary Figure 5. Gene-set enrichment analysis (GSEA) of 22 immune pathways across 15 cell types of nasal tissues.** The gradient color bar shows the normalized enrichment score (NES) score. NES score > 0 indicates the immune pathway significantly enriched in upper-regulated genes. NES score < 0 indicates the immune pathway significantly enriched in down-regulated genes. Black dots denote the FDR < 0.05.

### Supplementary Figure 6

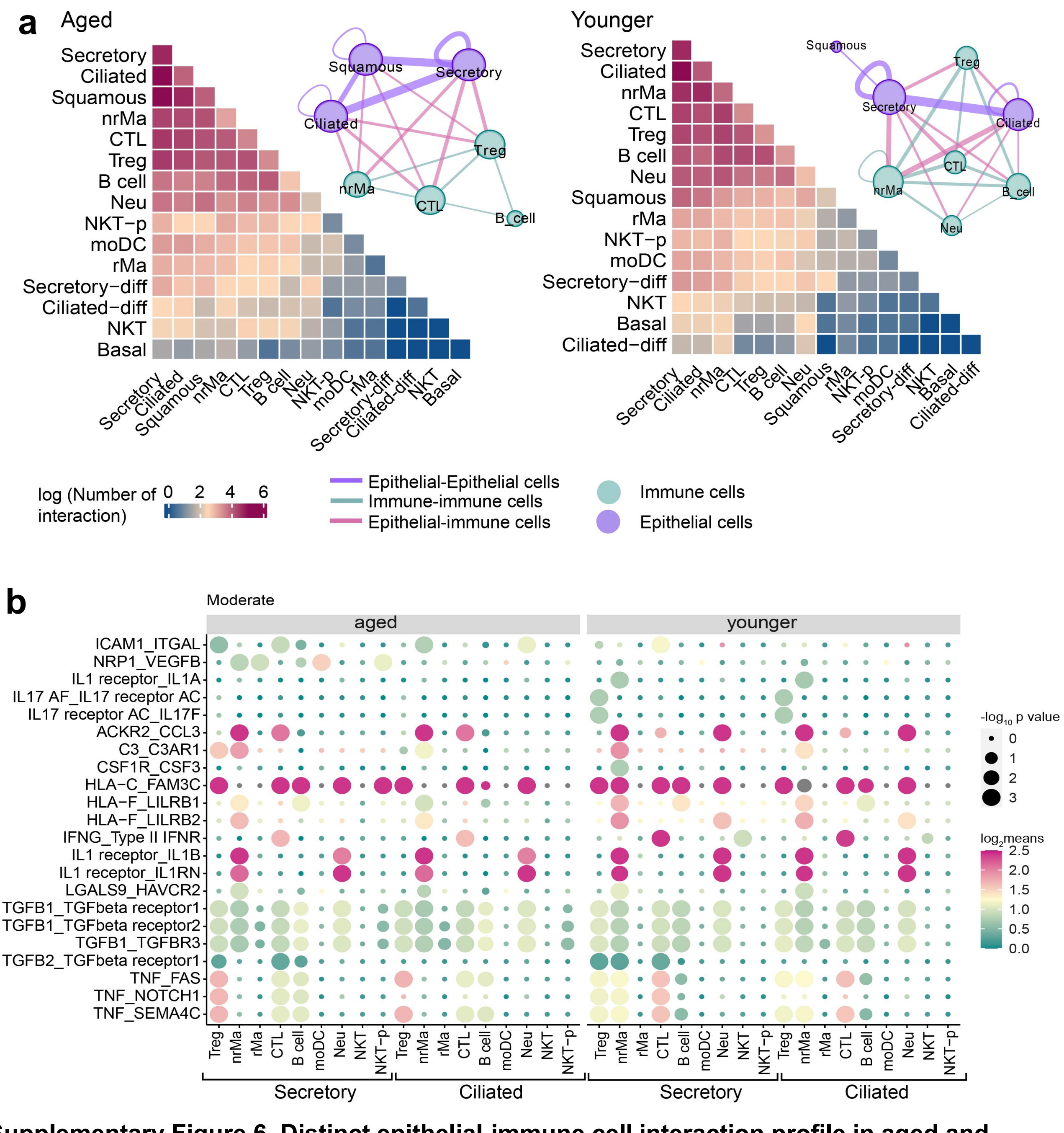


**Supplementary Figure 6. Distinct epithelial-immune cell interaction profile in aged and younger patients with moderate COVID-19. a,** Heatmap show the total log-scaled interaction number between epithelial-immune cells in moderate COVID-19 disease. Aged group, n= 3 patients, younger group, n = 4 patients. The cell-cell interaction network depicted all cell pairs which the number of cell-cell interaction > 50. Edge size denotes the number of interactions between two cell types. Different colors indicate the immune or epithelial cell types. **b,** Dot plot showing the significant ligand-receptor interactions between epithelial-immune cell interaction in moderate COVID-19 disease. The circle size indicated -log10-scaled p values by permutation test. Gradient color bar shows the log2-scaled means of average expression of interacted cell pair.

#### Supplementary Figure 7

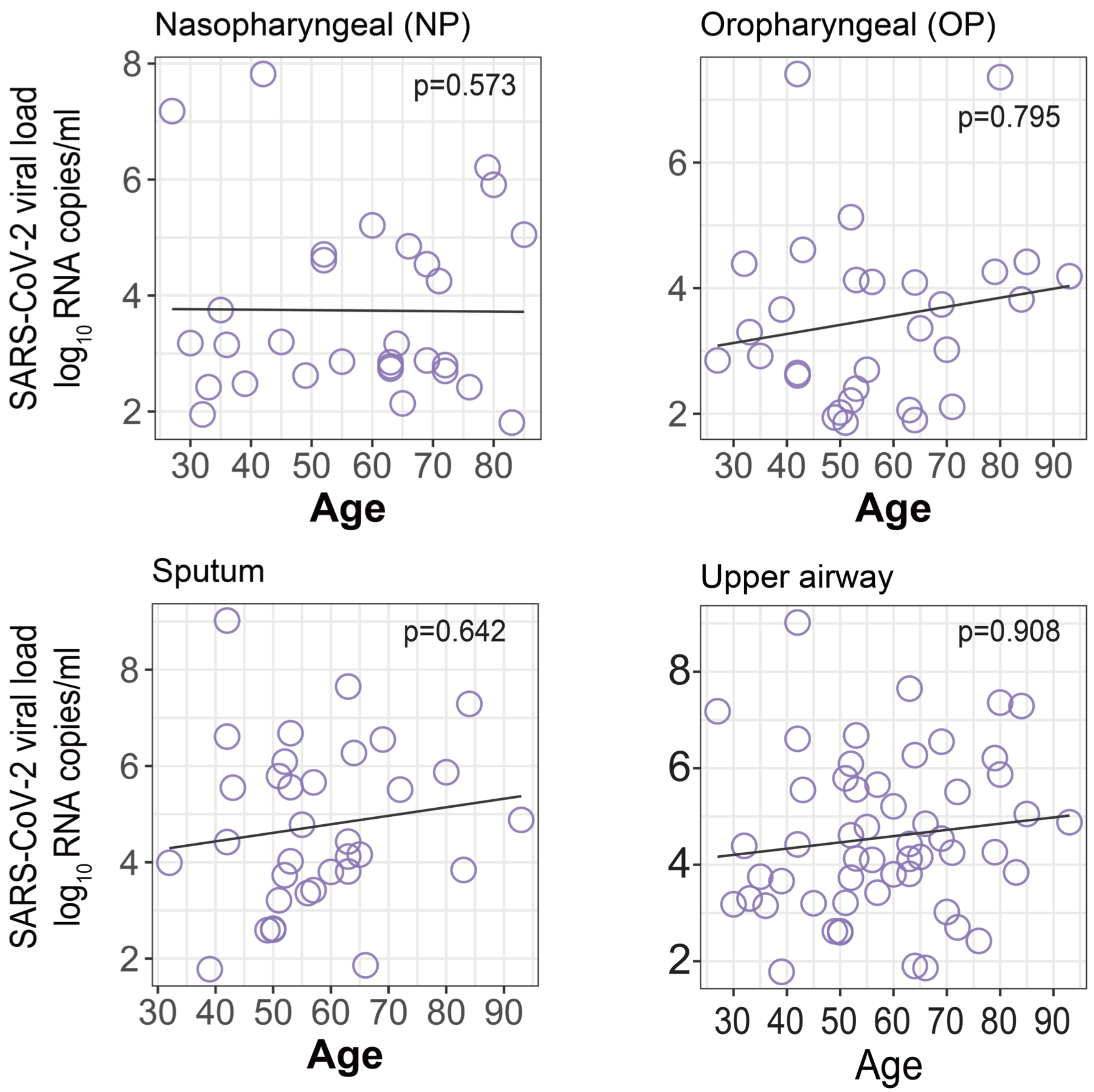


**Supplementary Figure 7. Correlation analysis between age and upper airway viral load**. The upper airway data from three sample source, oropharyngeal swab, nasopharyngeal and sputum.
